# The clinical pharmacology of tafenoquine in the radical cure of *Plasmodium vivax* malaria: an individual patient data meta-analysis

**DOI:** 10.1101/2022.09.12.22279840

**Authors:** James A Watson, Robert J Commons, Joel Tarning, Julie A Simpson, Alejandro Llanos Cuentas, Marcus VG Lacerda, Justin A Green, Gavin CKW Koh, Cindy S Chu, François H Nosten, Ric N Price, Nicholas PJ Day, Nicholas J White

## Abstract

**Background:** Tafenoquine is a newly licensed antimalarial drug for the radical cure of *Plasmodium vivax* malaria. The mechanism of action and the optimal dosing - a balance between haemolytic risk and radical cure efficacy - are uncertain.

**Methods:** We pooled individual data from 1102 patients with acute vivax malaria and 72 healthy volunteers (all with >70% of the population median whole blood glucose-6-phosphate dehydrogenase [G6PD] activity) who were studied in the pre-registration trials of tafenoquine. Plasma tafenoquine concentrations were analysed under a population pharmacokinetic model. A series of Bayesian logistic and Emax dose-response models were fitted to the primary outcome of any *P. vivax* recurrence within 4 months. Acute vomiting and haemolysis were secondary outcomes. The key exposure variables were: the mg/kg tafenoquine dose; pharmacokinetic summaries of parent compound exposure (AUC_[0,*∞*)_, C_max_) and metabolism (terminal elimination half-life); and day 7 methaemoglobin concentration (%) as a measure of oxidative activity.

**Results:** Tafenoquine dose (mg/kg) was a major determinant of recurrence (odds ratio [OR]: 0.70 per mg/kg increase, 95% credible interval [CI]: 0.65-0.75). After adjustment for dose, the tafenoquine terminal elimination half-life (OR: 1.15 per day increase, 95% CI: 1.06-1.25), and the day 7 methaemoglobin concentration (OR: 0.81 per absolute percentage point increase, 95% CI: 0.65 to 0.99), but not the parent compound exposure, were also associated with recurrence. Under the Emax we estimate that the currently recommended 300mg dose in a 60kg adult (≈5mg/kg) results in approximately 70% of the maximal obtainable hypnozoiticidal effect. Increasing the dose to 7.5mg/kg (i.e. 450mg) would result in 90% reduction in the risk of *P. vivax* recurrence. Tafenoquine was well tolerated. No patients had severe haemolysis. Acute vomiting was not dose-related. In patients with normal G6PD enzyme concentrations, tafenoquine dose was associated with minor post-treatment haemoglobin reductions on days 2 or 3 (0.02 g/dL per mg/kg increase [95% CI: 0.04 to 0.00]).

**Conclusions and interpretation:** The currently recommended 300mg adult dose for radical cure of vivax malaria has suboptimal efficacy. Increasing to 450mg is predicted to increase radical cure rates substantially. The production of oxidative metabolites is central to tafenoquine’s hypnozoiticidal efficacy. Clinical trials of higher tafenoquine doses are now needed to characterise efficacy, safety and tolerability.

## Introduction

Tafenoquine is a newly licensed 8-aminoquinoline antimalarial drug for the radical treatment of relapsing *Plasmodium vivax* and *Plasmodium ovale* malarias; the first for 70 years. Tafenoquine is eliminated slowly and can be given as a single dose for radical cure or as weekly chemoprophylaxis [1, 2]. This provides a substantial operational advantage over the currently recommended radical treatment, primaquine, which is eliminated rapidly and needs to be given at least once daily for 7-14 days [3, 4]. Primaquine is metabolised rapidly to carboxyprimaquine (considered pharmacologically inert) and a series of reactive oxidative intermediates which are thought to be responsible for both antimalarial activity and haemolytic toxicity [5, 6]. The main limitation to use of the antimalarial 8-aminoquinolines is oxidative haemolysis. This can be life-threatening in glucose-6-phosphate dehydrogenase (G6PD) deficiency, which is very common in malaria endemic areas [7]. The optimum 8-aminoquinoline dose is therefore a balance between benefits (applicable to all treated patients) and risks (borne largely by the subgroup of patients with G6PD deficiency). Tafenoquine has been engineered to be much more stable than primaquine. The contribution of oxidative metabolites to the pharmacological activity of tafenoquine is unclear [8]. Indeed, the precise mechanism of action of the 8-aminoquinoline antimalarials in killing hypnozoites remains unknown.

Following a phase 2b dose-ranging trial, a single 300mg dose of tafenoquine (approximately 5mg base/kg) was chosen for phase 3 evaluation in adults [9]. The principal dose-limiting concern at the time was the haemolytic potential of tafenoquine in G6PD deficient heterozygote females [10], who often test as “normal” in G6PD deficiency rapid screens. The phase 3 tafenoquine registration trials overall were interpreted as showing approximate equivalence of 300mg tafenoquine compared with lower dose primaquine (15mg base daily for 14 days), although the pre-specified non-inferiority margin was not met, and in Southeast Asia, tafenoquine efficacy was significantly inferior to primaquine [1]. *P. vivax* parasites from Southeast Asia and Oceania are considered to require higher 8-aminoquinoline doses than elsewhere. The World Health Organization recommends a total primaquine dose of 420mg base (30mg daily) in these regions compared with 210mg (15mg daily) in other endemic regions [11]. Thus, in the Southeast Asian region, the 300mg adult dose of tafenoquine was inferior to a sub-optimal primaquine radical cure regimen, although the sample size was small (43 patients randomised to low-dose primaquine, and 73 randomised to tafenoquine 300mg). In the recently reported INSPECTOR trial, 300mg single dose tafenoquine was associated with very poor radical curative efficacy in Indonesian soldiers returning from Papua, Indonesia, an area of high malaria transmission [12]. Several hypotheses for these disappointing results have been proposed including underdosing [13] and an antagonistic interaction between dihydroartemisinin-piperaquine (DHA-PQP) and tafenoquine. Although a single dose tafenoquine recommendation for all adults has practical advantages, it results in substantial mg/kg variations in drug exposure. Taken together, it seems likely that the currently recommended adult dose of tafenoquine 300mg is inferior to optimal primaquine regimens in preventing relapses of vivax malaria in all endemic regions [13, 14].

The 8-aminoquinoline drugs are considered to be pro-drugs, although there has been some debate about this for tafenoquine [8]. For primaquine, plasma concentrations of the parent compound and the inactive carboxy metabolite can be measured readily, but characterisation of the active metabolites, or their biotransformed derivatives, has proven challenging. Tafenoquine has a terminal elimination half-life of about 15 days compared with 5 hours for primaquine, so capturing transient active metabolites is even more difficult. The current inability to measure the biologically active moiety impedes pharmacometric dose optimisation trials. We have suggested that increases in red cell methaemoglobin (MetHb) production following 8-aminoquinoline administration could be used as an *in-vivo* pharmacodynamic proxy of oxidative antimalarial activity [6]. MetHb is an oxidation product in which the ferrous iron (Fe^2+^) of haem has been oxidised to the ferric form (Fe^3+^). Data from over 500 *P. vivax* malaria patients given high-dose primaquine radical cure suggested that increases in day 7 blood methaemoglobin concentrations (approximately the day of peak methaemoglobinaemia in a daily primaquine radical cure regimen) were associated with greater radical curative efficacy [15].

To guide optimal tafenoquine dosing and to identify factors associated with greater radical cure efficacy and haemolysis, we analysed the relationship between tafenoquine dose, pharmacokinetics, MetHb production, haemoglobin reduction and recurrence of vivax malaria using data from the three tafenoquine registration trials conducted in patients with acute vivax malaria (DETECTIVE phase 2b, DETECTIVE phase 3, and GATHER) [1, 2, 9]. Rich sampling data from a phase 1 study of tafenoquine pharmacokinetics in healthy volunteers [16] were pooled with the patient data to develop a population pharmacokinetic model of tafenoquine and thus characterise individual patient tafenoquine exposure and elimination. Our results provide important insights into the pharmacodynamics of tafenoquine, notably the central importance of oxidative metabolites. From this we show that the currently recommended dose of tafenoquine has sub-optimal efficacy, and we propose evaluation of a new 50% higher dose that could reduce vivax malaria recurrences substantially with little predicted increase in the risk of severe adverse events.

## Methods

This study was a meta-analysis of individual volunteer and patient data gathered during the detailed pre-registration studies of tafenoquine. The flow diagram is shown in Supplementary Figure S1 shows how the different analysis data sets were constructed. The results of all four studies included have been published previously [1, 2, 9, 16]. Our analyses do not include data from the TEACH trial (single arm paediatric trial of weight based tafenoquine doses in symptomatic *P. vivax* [17]) or the INSPECTOR trial (returning soldiers [12]) as the individual patient data from these studies are not yet available to independent researchers.

### Ethical review

Anonymised individual patient data were obtained via ClinicalStudyDataRequest.com following approval of a research proposal from the Independent Review Panel. Re-use of existing, appropriately anonymized, human data does not require ethical approval under the Oxford Tropical Research Ethics Committee regulations (OxTREC).

### Data

In all studies, healthy volunteers and patients were required to give written, informed consent. All patients and volunteers were screened for G6PD deficiency. The patients were non-pregnant adults with uncomplicated *P. vivax* malaria and greater than 70% of the population median G6PD enzyme activity. The studies were approved by local review boards/ethics committees and were conducted in accordance with the Declaration of Helsinki and Good Clinical Practice guidelines.

We pooled clinical efficacy, laboratory, and pharmacokinetic data from four studies in adults: the phase 2b dose-ranging trial DETECTIVE part 1 (NCT01376167 [9]); the phase 3 confirmatory trial DETECTIVE part 2 (NCT01376167 [2]); the phase 3 safety trial GATHER (NCT02216123 [1]); and a phase 1 pharmacokinetic study which assessed drug-drug interactions in healthy volunteers (NCT02184637, laboratory and pharmacokinetic data only [16]).

#### Drug administration

In all studies tafenoquine was administered orally as a single dose. In the DETECTIVE and GATHER trials, tafenoquine was administered in combination with the standard chloroquine malaria treatment regimen for adults (1500mg base given over 2-3 days). In these trials primaquine, 15mg daily given for 14 days, was the comparator. The phase 1 study was a drug-drug interaction study, with volunteers randomised to either tafenoquine alone or tafenoquine in combination with DHA-PQP or artemether-lumefantrine (AL), the standard antimalarial treatment regimens in both cases. In DETECTIVE part 1, tafenoquine was administered as a capsule, whereas in the other studies it was administered as a tablet. Each capsule or tablet contained 188.2mg tafenoquine succinate equivalent to 150mg of tafenoquine base. The methods used for the measurement of plasma tafenoquine concentrations have been published previously [18].

### Causal model of tafenoquine radical cure

The overall structure of the analysis is guided by the hypothetical simplified causal model shown in Figure 1. Under this model, the relationship between plasma tafenoquine concentrations and its radical curative activity is mediated by the production of oxidative intermediates (i.e. active metabolites). These are probably produced largely within hepatocytes, but also have systemic activity. These metabolites drive not only the efficacy of the drug but also the main adverse effects, notably haemolysis and the conversion of intraerythrocytic haemoglobin to MetHb. Thus, as MetHb production and haemolysis both result from the production of active oxidative metabolites, they could be proxy markers of radical curative efficacy [6].

**Figure 1:**
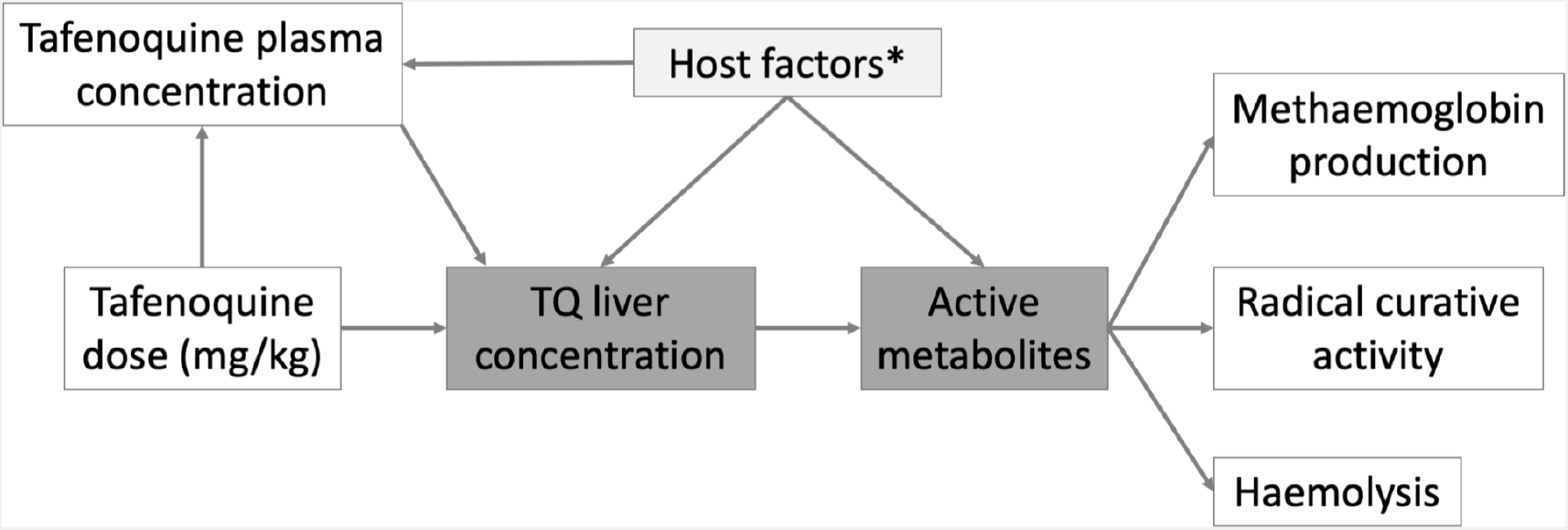
Hypothetical causal model of the clinical pharmacology of tafenoquine for the radical treatment of *P. vivax* malaria. The dark grey shaded boxes represent non-observables; lighter shaded grey represents partially observable measures. *Potential host factors include genetic factors such as *CYP2D6* polymorphisms; age related enzyme maturation; drug-drug interactions.

### Population pharmacokinetic modelling

Tafenoquine pharmacokinetics have been described in detail previously [18]. Tafenoquine is a slowly eliminated 8-aminoquinoline antimalarial drug with a terminal elimination half-life of around 15 days. The metabolites have not been well characterised. Tafenoquine pharmacokinetics have been described previously by a two-compartment pharmacokinetic (PK) model (i.e. bi-exponential decay model) with allometric scaling of clearance and volume parameters [18]. We inspected visually all individual drug level profiles and excluded 10 concentrations from 8 patients which were substantial outliers. A total of 15 malaria patients vomited within one hour of drug administration. The protocol for all three studies indicated that these patients should be re-dosed at the same dose. One patient was not re-dosed and had very low tafenoquine levels during follow-up. We removed this patient from all analyses as it is difficult to estimate the amount of drug ingested. For the remaining 14 patients who vomited and were re-dosed, time since dose was calculated as the time since the second dose (in some patients this was the next day). All available tafenoquine concentrations were transformed into their natural logarithms and characterised using nonlinear mixed-effects modelling in the software NONMEM v7.4 (Icon Development Solution, Ellicott City, MD). A total of 2.56% of observed tafenoquine concentrations were measured to be below the limit of quantification and subsequently replaced with a value equal to half of the limit of quantification [19]. One-, two- and three-compartment dispositional models were evaluated, as well as different absorption models (i.e. first-order absorption and a fixed number of transit absorption compartments). Inter-individual variability was implemented as an exponential function of all structural PK parameters. The first-order conditional estimation method with interactions (FOCE-I) and subroutine ADVAN5 TRANS1 were used throughout the study. Body weight was implemented as a fixed allometric function on clearance parameters (exponent of 0.75) and as a linear function on volume parameters (exponent of 1.0). Additional, biologically relevant, covariates (i.e. age, sex, vomiting, tafenoquine formulation and patient vs healthy volunteers) were evaluated on all PK parameters using a linearized stepwise addition (p<0.01) and backward elimination (p<0.001) covariate approach. Statistically significant covariates retained in the backward elimination step were evaluated further for clinical significance and retained only if the range of individual covariate values resulted in a greater than +/-10% effect compared with the population mean value.

Potential model misspecification and systematic errors were evaluated by basic goodness-of-fit diagnostics. Model robustness and parameter confidence intervals were evaluated by a sampling-important-resampling (SIR) procedure [20]. Predictive performance of the final model was illustrated by prediction-corrected visual predictive checks (n = 2,000) [21]. The 5th, 50th, and 95th percentiles of the observed concentrations were overlaid with the 95% confidence intervals of each simulated percentile to detect model bias. Under the population mixed-effects model we estimated for each healthy volunteer and each patient in the three efficacy studies: (i) the area under the plasma concentration time curve (AUC_[0,*∞*_); (ii) the maximum peak concentration (*C*_*max*_); and the terminal elimination half-life (t1/2). AUC_[0,*∞*_ and *C*_max_ are summary statistics of the individual’s exposure to the parent compound; whereas *t*_1*/*2_ is a summary statistic of the metabolism of the parent compound (tafenoquine is eliminated via biliary excretion with enterohepatic recirculation, but is not eliminated in the urine [22]).

For the terminal elimination half-life, multiple patients had estimated values greater than 30 days. On examination, the NONMEM model estimates of the terminal elimination half-life were not robust with respect to the model structure. In order to derive robust half-life estimates, we fit a robust (using a student-*t* likelihood [23]) Bayesian hierarchical linear model to the observed log-concentrations post day 7 (terminal phase; random intercept and slope). Patients had a median of 3 values post day 7 (IQR: 2-3). The half-life is then defined as −log(2)*/β*_*i*_ where *β*_*i*_ is the individual slope estimate for patient *i*. Values below the lower limit of quantification were treated as left-censored observations with censoring value equal to log(2ng/ml). The model included an interaction term between time and the log patient weight (clearance was associated with weight); and a dose (mg/kg) dependent intercept term. We then calculated the weight-adjusted terminal elimination half-life by subtracting the effect of weight estimated under a linear model.

The majority of patients in the three efficacy trials had between 5 and 6 observed plasma tafenoquine samples above the lower limit of quantification (Supplementary Figure S3). We note that for patients with few samples (18 patients had fewer than 4 samples) the PK summary statistics (AUC_[0,*∞*_, *C*_max_, *t*_1*/*2_) will be shrunk towards the population mean values.

### Methaemoglobin

All patients in the phase 1 trial and the three efficacy trials had frequent blood MetHb measurements taken using non-invasive signal extraction pulse CO-oximeter handheld machines. For the three efficacy trials, we excluded measurements taken at or after recurrence of vivax malaria. Because erroneous readings from the handheld oximeters can occur, we inspected visually the longitudinal MetHb curves for each patient. This visual check showed that all patients from one study site in Thailand had highly implausible measurements, most likely as a result of transcribing the carboxyhaemoglobin and not the MetHb value. All patients from this site were excluded from the MetHb analysis (*n*=91), resulting in a total of 9632 MetHb measurements across 746 *P. vivax* infected patients and 72 healthy volunteers. Between the time of first dose (defined as the time of the first chloroquine or DHA-PQP dose for the patients who did not receive tafenoquine, and the time of the first tafenoquine dose for those who did) and day 20, there were a total of 5816 measurements, with a median (range) of 7 measurements per patient (1 to 12).

Following our previous analysis, which examined the relationship between MetHb concentrations and *P. vivax* recurrence after high-dose primaquine [15], we summarised MetHb production using the day 7 value (approximately the day of the peak value). In patients who did not have a measurement on day 7, we used linear interpolation to estimate a day 7 value.

### Radical curative efficacy

#### Rationale for the primary endpoint

The primary endpoint for the main efficacy analyses was any *P. vivax* recurrence within 4 months. We chose the binary endpoint of any recurrence rather than the time to first recurrence because tafenoquine has both asexual stage antimalarial activity [24] and hypnozoiticidal activity. Thus, an association between the mg/kg tafenoquine dose and the time to recurrence could, in theory, result from longer periods of post-treatment prophylaxis (i.e. suppression of blood stage multiplication) with higher mg/kg doses. Plasma concentrations above a plausible minimum inhibitory concentration for the asexual activity persist for up to three months [25]. Therefore, if relapses are delayed by the post-treatment prophylactic levels, we would expect to detect them before a 4-month cut-off, and definitely before a 6-month cutoff (ignoring long latency relapse, which will, in general, not be observed with 6 months follow-up). The choice of a particular cut-off for follow-up is thus a trade-off between sensitivity and specificity. Longer follow-up will increase the sensitivity of the endpoint (include more relapses) but decrease specificity (include more reinfections).

We chose a 4-month cut off for the primary endpoint for the following two reasons:

- *A priori* we think that the 4-month endpoint will have very high sensitivity. This is >95% based on the data from the INSPECTOR trial. The INSPECTOR trial showed that in returning Indonesian soldiers, in whom reinfections were not possible, over 95% of all relapses following treatment with DHA-PQP plus tafenoquine occurred within 4 months [12]. Very large field trials in patients given no radical cure and with multiple ACT partner drugs confirm the generalisability of this finding [4, 26]. For example, in the study by Chu *et al*, 90% of relapses were estimated to have occurred by 9 weeks [26]. Extending the follow-up to 6 months will have minimal impact on sensitivity but will reduce specificity (by an amount that is directly proportional to local transmission intensity).
- The 4-month endpoint has a much lower proportion of loss-to-follow-up (i.e. incompletely captured data) compared to the 6-month endpoint. After excluding patients whose last visit was before day 31 (a total of 27 patients who are non-informative due to chloroquine post-treatment prophylaxis: no recurrences are expected in the first month [4]), only 12 out of the 1073 remaining patients did not have a 4 month follow-up visit (1.1%), compared to 98 out of 1073 who did not have 6 month follow-up visit (9.1%).

The main limitation of the binary endpoint of ‘any recurrence’ relates to the slightly longer post-prophylactic periods in the tafenoquine treated patients: their at-risk period for reinfection is shorter. All patients were treated with chloroquine after which the first detected recurrence (either reinfection or relapse) occurs approximately 1.5 months later [27]. High doses of tafenoquine could plausibly delay recurrence for another 2 weeks [25]. However, the post-prophylactic effect of both drugs most likely delays rather than suppresses reinfection occurring a few weeks after treatment [28]. Thus, bias resulting from a greater number of reinfections in the placebo versus tafenoquine treated groups is likely to be small.

#### Efficacy models

We explored the association between the following variables and the odds of *P. vivax* recurrence (the 12 patients with incomplete follow-up were coded as ‘successes’, i.e. no recurrent infection):

1. The mg/kg dose of tafenoquine, based on the recorded patient body weight (patients who received primaquine in comparator arms are included with an additional indicator variable multiplied by the mg/kg dose of primaquine);
2. The AUC_[0,*∞*)_ of the plasma tafenoquine concentration (only patients who received tafenoquine), estimated from the population PK model;
3. The tafenoquine *C*_max_ (only patients who received tafenoquine), estimated from the population PK model;
4. The tafenoquine terminal elimination half-life *t*_1*/*2_ (only patients who received tafenoquine), estimated from a robust linear model fit to the terminal phase data (post day 7), and with adjustment for tafenoquine dose;
5. The day 7 MetHb in a model adjusted for the mg/kg dose of tafenoquine (only patients who received tafenoquine);

For all predictors we fitted Bayesian mixed effects logistic regression models (random intercepts for study and study site), with adjustment for day 0 parasitaemia (log transformed). Logistic regression allows inference of the odds-ratio for recurrence for the predictive variable of interest, assuming a linear relationship between the predictive variable and log-odds for recurrence.

For the mg/kg dose (the primary predictor of interest), the dose-response relationship was then assessed under a Bayesian E_max_ model, where the dose-dependent logit probability of recurrence is defined as:

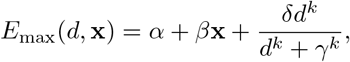

where *d* ≥ 0 is the tafenoquine dose (mg/kg) and **x** is a vector of additional covariates (for example, in the analyses which include data from primaquine treated patients, **x** would include the primaquine mg/kg dose); *α* is the intercept on the logit scale; *δ* determines the maximal tafenoquine effect (on the logit scale), i.e. the minimal logit probability of recurrence (an asymptote for very large doses); *k* is the slope coefficient; *γ* is the half-maximal effect (on the logit scale). The E_max_ model was also fitted with random intercepts for study and study site.

The E_max_ model is the most biologically plausible model for the pharmacodynamic effect of tafenoquine as estimated in these studies. No dose of tafenoquine is expected to reduce the number of recurrences by four months completely to zero as some of the recurrences will be reinfections.

#### *CYP2D6* polymorphisms

*CYP2D6* genotyping data were available for patients enrolled in DETECTIVE part 2 and GATHER. We mapped *CYP2D6* diplotypes to an activity score using the standard scoring system [29] (note that under the updated scoring system the *10 allele has a score of 0.25 instead of 0.5). The activity score cannot be used as a linear predictor (e.g. a score of 1 cannot be interpreted as twice the metabolic activity of a score of 0.5). For this reason, to assess association with recurrence, we dichotomised the activity scores into poor metabolisers (≤0.5) versus normal/extensive metabolisers (>0.5) [29].

#### Comparison of predictors of recurrence

To determine the main predictors of recurrence, we fitted a multivariable penalised Bayesian logistic regression model. This included the following variables as predictors: the tafenoquine mg/kg dose; the plasma tafenoquine AUC_[0,*∞*)_; the plasma tafenoquine *C*_max_; the day 7 MetHb concentration (%); the terminal elimination half-life *t*_1*/*2_; and the baseline parasite density. The model included random intercept terms for each study and study site. Each fixed effect predictor was scaled to have mean 0 and standard deviation 1. To avoid convergence issues resulting from the co-linearity of the predictors, the prior on all non-hierarchical coefficients was set to a Normal(0,0.5). This prior effectively shrinks the regression coefficients towards zero. It can be interpreted as follows: plausible odds-ratios for recurrence for one standard deviation change for each of the fixed effect predictors lie approximately between 0.38 and 2.7).

#### Sensitivity analyses

In *a priori* sensitivity analyses, all recurrences up until 6 months were included as both binary and time to event outcomes. The time to first recurrence was defined as a *P. vivax* episode between day 7 and day 180 with right censoring at the last recorded visit. We also re-ran the same models in patients who received a 300mg single dose of tafenoquine only (i.e. the current recommended treatment).

### Tolerability and safety analyses

For tolerability, acute vomiting was defined as vomiting within 1 hour of tafenoquine dosing and for safety three definitions of severe haemolysis were assessed:

- Relative fall in haemoglobin by more than 25% to <7g/dL
- Absolute fall in haemoglobin greater than 5 g/dL
- Any haemoglobin fall to <5 g/dL

Tolerability and safety events were categorised according to tafenoquine treatment and doses (no tafenoquine, <3.75mg/kg tafenoquine, [3.75,6.25) mg/kg tafenoquine, [6.25,8.75) mg/kg tafenoquine, ≥8.75mg/kg tafenoquine). The change in haemoglobin concentration from day 0 to the minimum on day 2 or 3, the expected day of nadir [30], was calculated to assess changes in haemoglobin associated with tafenoquine dose. Mixed effects models were used to explore the association between tafenoquine mg/kg dose, day 7 MetHb concentration, AUC_[0,*∞*)_, C_max_, and *t*_1*/*2_ adjusting for age, sex, day 0 parasite density and day 0 haemoglobin, with random effects for study and site.

### Statistical analyses

All analyses were done using R version 4.0.2 and Stata v17.0. Bayesian mixed-effects logistic regression models were fitted using the *rstanarm* package using default prior distributions. The Bayesian E_max_ models were coded in stan and fitted using the *rstan* interface. Survival models were fitted in Stata. Population pharmacokinetic models were fitted using NONMEM v7.4. Terminal elimination half-lives were estimated using the *brms* package based on stan [31].

### Data availability

Data can be accessed by submitting an application via www.ClinicalStudyDataRequest.com using the following study codes: GSK-TAF112582; GSK-200951; GSK-TAF116564.

### Code availability

All analysis code is available at the following github repository: https://github.com/jwatowatson/Tafenoquine-efficacy.

## Results

### Weight-adjusted radical cure efficacy of single-dose tafenoquine

Efficacy data were pooled from 1073 patients with acute *P. vivax* malaria who were recruited in nine countries (Table 1) between October 2010 and May 2017 (Supplementary Figure S1). All patients were treated with a standard dose regimen of chloroquine (1500mg base over 3 days) and were randomised to either single dose tafenoquine (varying doses), low-dose primaquine (15mg base daily for 14 days), or to no 8-aminoquinoline treatment. In the patients who received tafenoquine (*n*=634), single doses ranged from 50mg to 600mg, with weight adjusted doses ranging from 0.5mg base/kg to 14.3mg/kg (i.e. a greater than 25-fold variation of weight adjusted dose). Figure 2 shows the Kaplan-Meier survival curves for the pooled data on time to first recurrence, grouped by tafenoquine dosing category, and compared with low-dose primaquine (i.e. adult dose 15mg/day for 14 days) and no radical cure regimen. Patient weight (and thus mg/kg tafenoquine dose) varied considerably across the nine countries, with the heaviest patients in Brazil (median weight 70kg) and the lightest patients in Ethiopia and the Philippines (median weight 53kg), see Supplementary Figure S4.

**Table 1:** Demographic, clinical and dosing information on tafenoquine and primaquine dosing groups for all patients included in the efficacy analyses. (*n* = 1073, see Supplementary Figure S1). Unless otherwise stated, for binary variables we show the total number (%); for continuous variables we show the mean (standard deviation). * Median (interquartile range).

| | No TQ<br>$n=182$ | PQ<br>$n=257$ | TQ<br><3.75<br>mg/kg<br>$n=169$ | TQ<br>[3.75,6.25)<br>mg/kg<br>$n=368$ | TQ<br>[6.25,8.75)<br>mg/kg<br>$n=54$ | TQ $\geq 8.75$<br>mg/kg<br>$n=43$ |
| --- | --- | --- | --- | --- | --- | --- |
| Recurrence at 4 months | 101 (55.5) | 57 (22.2) | 57 (33.7) | 79 (21.5) | 4 (7.4) | 0 (0) |
| Age (years) | 35 (14.1) | 36 (14.4) | 37 (13.4) | 36 (14.2) | 36 (17.2) | 33 (14.4) |
| Female | 50 (27) | 77 (30) | 43 (25) | 95 (26) | 24 (44) | 9 (21) |
| Weight (kg) | 62 (12.0) | 63 (12.1) | 72 (20.1) | 63 (8.7) | 51 (13.3) | 57 (7.3) |
| Baseline para-sitaemia (/uL)* | 5470 (2173 - 11856) | 4697 (1712 - 10430) | 4320 (1447 - 9456) | 4174 (1431 - 10101) | 5507 (1961 - 12111) | 6143 (1692 - 13313) |
| Haemoglobin day 0 (g/dL) | 12.9 (1.5) | 13.0 (1.6) | 12.9 (1.8) | 13.2 (1.6) | 12.3 (1.8) | 12.6 (1.8) |
| *Country* |  |  |  |  |  |  |
| Brazil | 55 (30) | 80 (31) | 61 (36) | 105 (29) | 6 (11) | 3 (7) |
| Cambodia | 10 (5) | 8 (3) | 0 (0) | 16 (4) | 2 (4) | 0 (0) |
| Colombia | 0 (0) | 5 (2) | 0 (0) | 11 (3) | 0 (0) | 0 (0) |
| Ethiopia | 14 (8) | 13 (5) | 0 (0) | 18 (5) | 7 (13) | 0 (0) |
| India | 9 (5) | 5 (2) | 21 (12) | 8 (2) | 4 (7) | 7 (16) |
| Peru | 62 (34) | 91 (35) | 56 (33) | 136 (37) | 21 (39) | 18 (42) |
| Philippines | 1 (1) | 2 (1) | 0 (0) | 3 (1) | 0 (0) | 0 (0) |
| Thailand | 31 (17) | 38 (15) | 31 (18) | 46 (12) | 10 (19) | 15 (35) |
| Vietnam | 0 (0) | 15 (6) | 0 (0) | 25 (7) | 4 (7) | 0 (0) |

**Figure 2:**
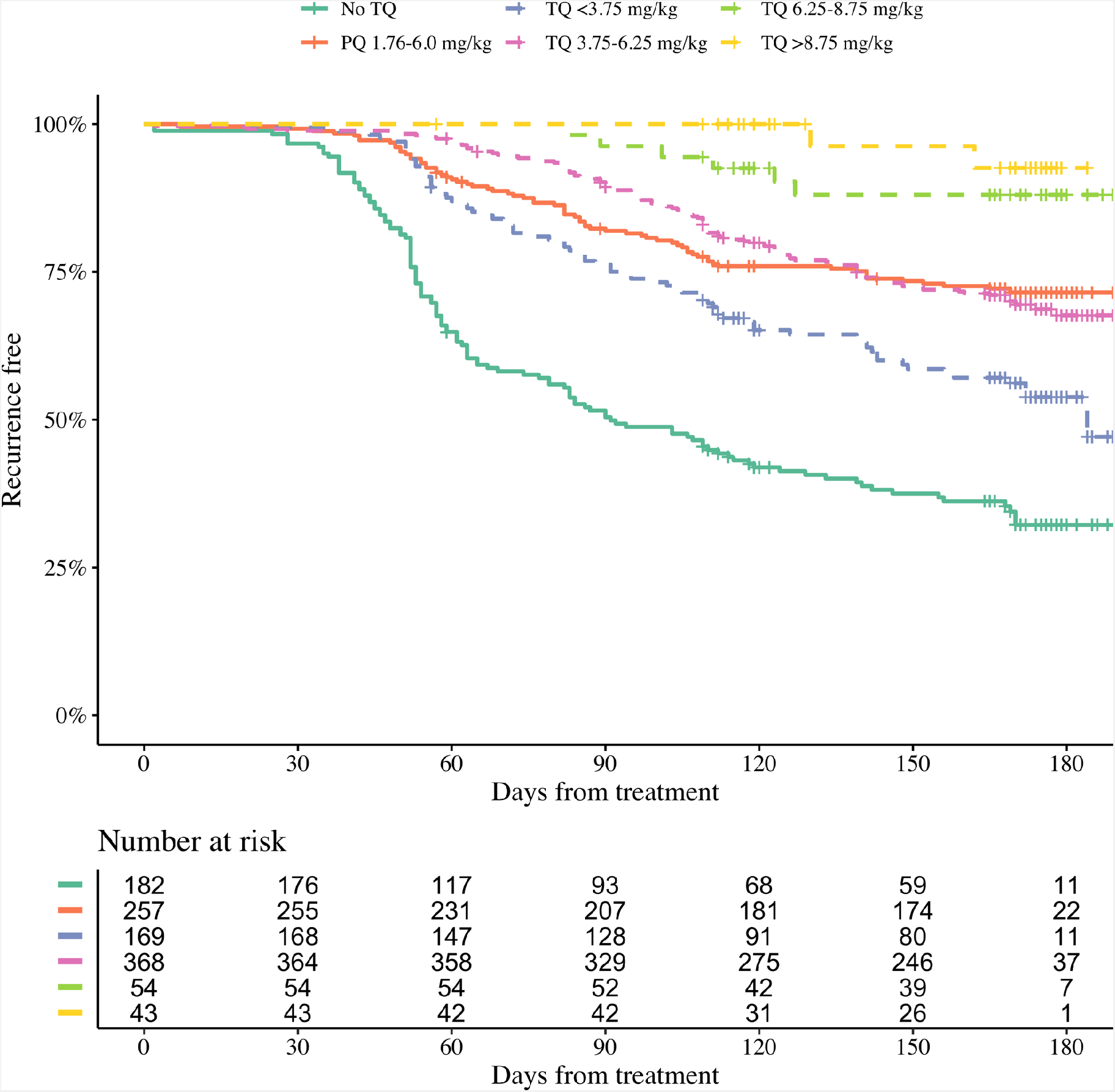
Kaplan-Meier survival curves for the time to first recurrence in 1073 *P. vivax* patients. Patients are grouped by tafenoquine mg/kg dosing category (dashed and dotted lines) versus low-dose primaquine (15mg base for 14 days) or no radical cure (thick lines). The vertical lines show the censoring times.

We first characterised the association between the mg/kg tafenoquine dose and the odds of recurrence using logistic regression. Under the model fit to all data (*n*=1073), each additional mg/kg of tafenoquine was associated with an odds ratio (OR) for recurrence within 4 months of 0.70 (95% CI: 0.65 to 0.76). In comparison, each additional mg/kg of primaquine was associated with an odds ratio of 0.62 (95% CI 0.55 to 0.71). The effect of tafenoquine mg/kg dose was similar in an analysis restricted to patients who received the currently recommended 300mg dose (*n*=469, OR=0.66; 95% CI 0.51 to 0.85). The association between tafenoquine mg/kg dose and recurrence was observed in all three geographic regions (Americas, Horn of Africa and Southeast Asia: Supplementary Figure S5), with near identical results when using any recurrence by 6 months as the endpoint or analysing by time to first recurrent event (Supplementary Figures S6 and S7). Further analysis under the E_max_ model indicated that the mg/kg dose-recurrence relationship was not linear on the log-odds scale. No recurrences by 4 months were observed in any patients in the largest dosing group (≥8.75, *n*=43). The E_max_ model estimated a slightly steeper dose-response curve compared to logistic regression fits (Supplementary Figure S8). Figure 3 shows the estimated dose-response curve for recurrence at 4 months under the E_max_ model. Taking 15mg/kg as the maximum tolerated dose of tafenoquine, a dose of approximately 7.5mg/kg achieves 90% of the effect achieved with 15mg/kg (Supplementary Figure S9). Near identical results were obtained for the 6 month endpoint (Supplementary Figure S10). Under the E_max_ model, the dose-response is steep around doses of approximately 2.5mg/kg to 5mg/kg. For example, increasing the dose from 3mg/kg to 4mg/kg reduces the mean estimated recurrence from 28.5% to 20.8%; increasing to 5mg/kg gives a mean recurrence of 14.2%.

**Figure 3:**
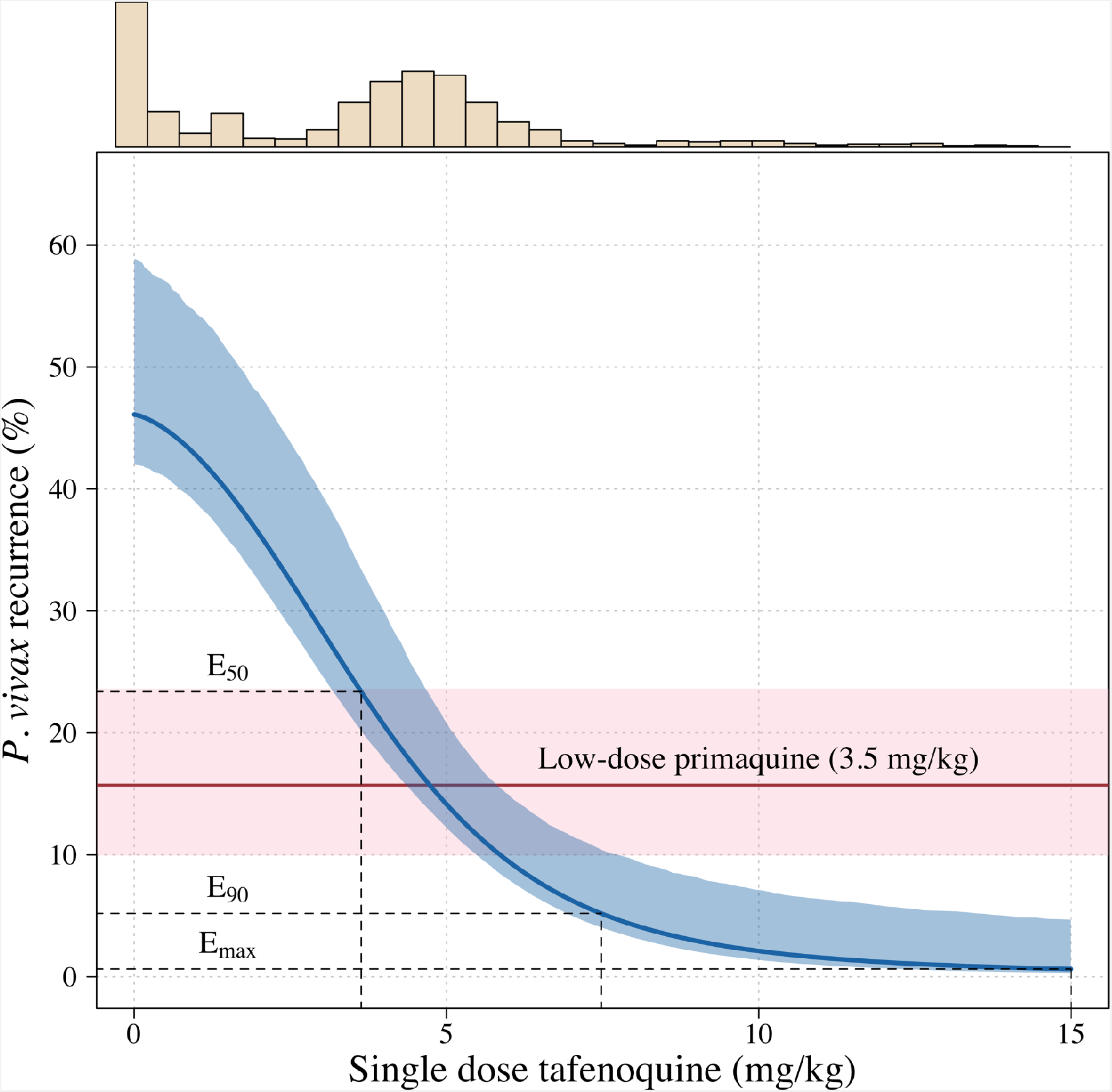
Tafenoquine mg/kg dose and the probability of a recurrence of *P. vivax* malaria within 4 months under the E_max_ model. Thick blue (shaded blue): mean probability of recurrence as a function of the tafenoquine mg/kg dose (95% CI); thick red (shaded pink): mean probability of *P. vivax* recurrence after 3.5mg/kg primaquine (95% CI). The distribution of administered tafenoquine mg/kg doses is shown by the histogram above (this includes patients who received no radical cure treatment, but for clarity does not include patients who received primaquine: *n*=816; range: 0 to 14.3mg/kg).

### Predicted efficacy of higher dose tafenoquine

Using the empirical weight distribution from the three efficacy trials, we estimate that a fixed 300mg dose in an adult would result in a mean recurrence proportion of 15.3% (95% CI: 9.6 to 21.7); whereas a 450mg fixed dose would result in a mean recurrence proportion of 6.2% (95% CI: 3.2 to 11.5). Given that approximately half the patients with no 8-aminoquinoline treatment had a recurrence, this implies that the 300mg fixed dose reduces recurrence rates by around 70%, whereas a 450mg fixed dose would reduce them by around 85%. Supplementary Figure S11 shows the expected distributions of individual efficacy (relative to all patients receiving 15mg/kg) for the 300mg versus 450mg fixed dose, based on the empirical distribution of weights in the three efficacy studies. This illustrates that small dose increments can result in substantial increases in radical cure efficacy. For the 450mg versus 300mg dose, this translates to a number needed to treat of 11 patients (95% CI 7 to 21) to prevent one recurrence of malaria (Supplementary Figure S12) with the greatest absolute benefit for patients weighing over 60kg (Supplementary Figure S13).

### Pharmacokinetic predictors of tafenoquine radical cure efficacy

We fitted mixed-effects population pharmacokinetic models to all available plasma tafenoquine levels (a total of 4499 tafenoquine plasma concentrations in 718 individuals: 72 healthy volunteers and 646 patients with symptomatic *P. vivax* malaria). The data were described adequately by a two-compartment elimination model, consistent with previous analyses of these data [18]. For each patient who received tafenoquine and who had at least one measurable plasma level recorded (*n*=646), drug exposures and metabolism were summarised by the total area under the plasma concentration time curve (AUC_[0,*∞*)_), the maximum plasma concentration (C_max_), and the terminal elimination half-life (*t*_1*/*2_).

Higher plasma tafenoquine AUC_[0,*∞*)_ values were associated with lower odds of recurrence at 4 months (OR: 0.61 [95% CI 0.47 to 0.77] for each standard deviation increase in AUC_[0,*∞*)_). However, this relationship was driven primarily by dose, i.e. the differences in AUC_[0,*∞*)_ between those who received lower (50mg or 100mg) versus higher (300mg or 600mg) doses (Supplementary Figure S14). In a model adjusted for tafenoquine mg/kg dose, the AUC_[0,*∞*)_ was not associated with recurrence at 4 months (OR: 0.89 per standard deviation change in AUC_[0,*∞*)_, 95% CI 0.64 to 1.23). The same pattern was seen for the C_max_: in a univariable model there was a strong association between C_max_ and recurrence, but in a model adjusted for tafenoquine mg/kg dose this association was no longer significant (OR: 1.01 for a doubling in C_max_, 95% CI 0.69 to 1.49). Thus dose administered rather than parent compound exposure was the main observed driver of therapeutic efficacy.

Assuming linear kinetics, the weight-adjusted *t*_1*/*2_ is, by definition, independent of the mg/kg dose. In contrast to exposure data (AUC_[0,*∞*)_ and C_max_), the individual tafenoquine weight-adjusted *t*_1*/*2_ estimates were associated with recurrence at 4 months. More rapid tafenoquine elimination (despite the lower parent drug exposure) was associated with lower odds of recurrence. In a model adjusted for tafenoquine mg/kg dose, the odds ratio for recurrence at 4 months for each day increase in *t*_1*/*2_ was 1.15 (95% CI 1.06 to 1.25).

### Methaemoglobin production and tafenoquine radical cure efficacy

Increases in blood MetHb concentrations following tafenoquine administration were highly correlated with the mg/kg dose (Figure 4 a-c). Each additional mg/kg was associated with a 19% (95% CI: 17% to 21%) increase in day 7 MetHb concentrations. As reported previously for high-dose primaquine [15], peak MetHb levels were observed around day 7 (Figure 4 a-b). Adding the day 7 MetHb concentration as a linear predictor in the logit-Emax model (i.e. in a model adjusted for the mg/kg dose of tafenoquine), each absolute percentage point increase in the day 7 MetHb concentration was associated with an odds ratio for recurrence of 0.81 (95% CI 0.65 to 0.99). Furthermore, consistent with this association between MetHb and vivax malaria recurrence (Figure 1), the day 7 MetHb levels were weakly inversely correlated with the tafenoquine weight-adjusted *t*_1*/*2_ values (Figure 4d; *ρ* = −0.1, p=0.05).

**Figure 4:**
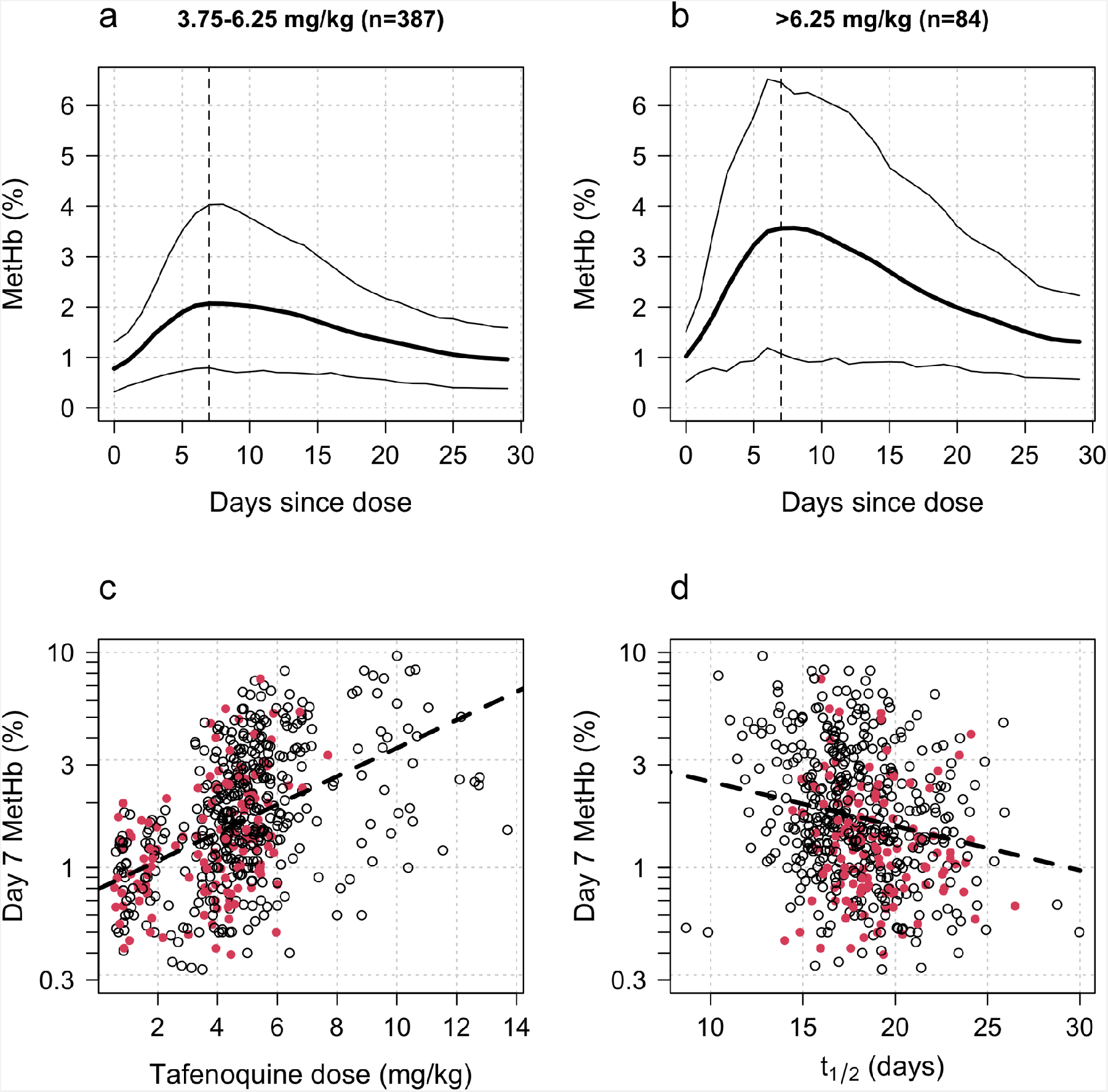
Tafenoquine dose, metabolism, and MetHb production. Panels a & b show the dose-dependent increases in blood MetHb concentrations expressed as a proportion (%) of the haemoglobin concentration following tafenoquine administration (mean: thick lines; 10th and 90th percentiles: thin lines). The approximate peak concentrations occur one week after receiving tafenoquine (shown by the vertical dashed lines). Panel c shows the relationship between the administered tafenoquine doses (mg/kg) and the interpolated MetHb concentrations on day 7 (log_10_ scale). Panel d shows the relationship between the estimated individual tafenoquine terminal elimination half-lives (*t*_1*/*2_) and the interpolated MetHb concentrations on day 7 (log_10_ scale). In panels c & d patients who had a recurrence within 4 months are shown by the filled red circles.

The high failure rates of tafenoquine plus DHA-PQP in the INSPECTOR study [12] raised concerns over drug-drug interactions between tafenoquine and partner schizontocidal drugs. We compared day 7 MetHb concentrations in patients and healthy volunteers as a function of the partner antimalarial drug (all *P. vivax* patients were given chloroquine, *n*=578; healthy volunteers received either no partner drug [*n*=24], DHA-PQP [*n*=24], or artemether-lumefantrine (AL) [*n*=24]), adjusting for the tafenoquine mg/kg dose (the major predictor of day 7 MetHb). Overall, after adjustment for the mg/kg dose, healthy volunteers had slightly lower day 7 methaemoglobinaemia compared to patients infected with *P. vivax* (15% lower, 95% CI: 0-34% p=0.02), Supplementary Figure S15. When comparing by randomised arm in the healthy volunteer trial, the largest observed difference was in those who received DHA-PQP in whom the day 7 MetHb was 23% lower (95% CI: 1.5 to 40%) compared with those volunteers who were not given a partner drug. However, it is not possible to differentiate between drug effects and disease effects when comparing malaria patients with healthy volunteers.

### *CYP2D6* polymorphisms

Cytochrome P450 2D6 (CYP2D6) is an enzyme that metabolises primaquine to a series of bioactive hydroxylated metabolites [5]. Genetic polymorphisms in *CYP2D6* resulting in loss of function are associated with reduced primaquine radical curative efficacy [32, 33]. A total of 716 patients had *CYP2D6* genotyping data, with a wide range of variants observed. The most common poor metaboliser alleles were *10 (10.5% allele frequency), *4 (5.8% allele frequency) and *5 (3.1% allele frequency). In the efficacy analysis population, of those randomised to tafenoquine, a total 34 (8.9%) patients had a predicted enzyme activity score of 0.5 or less (poor metabolisers) [29]; 67% (23/34) of these were in patients enrolled in Southeast Asia where *10 has high allele frequencies. Consistent with the previous analysis of these data [34] there was no clear association between the activity score and recurrence at 4 months following tafenoquine (OR for recurrence: 0.9 [95% CI 0.4-2.2] for poor versus normal/extensive metabolisers), although statistical power was low as few patients had low activity scores.

### Tolerability and safety

In total 2.3% (15/651) of patients vomited within one hour of oral tafenoquine administration. There was no clear association with the mg/kg dose (OR for vomiting per mg/kg increase: 1.08 [95% CI: 0.88 to 1.32]). Severe haemolytic events were rare. One patient treated with tafenoquine had a greater than 5g/dL fall in Hb, but no patients had a greater than 25% fall in Hb to <7g/dL, or a fall to less than 5g/dL. In a mixed effects linear regression model each additional mg/kg of tafenoquine was associated with a haemoglobin change at day 2 or 3 of -0.02g/dL (95% CI: -0.04 to 0.00; *p* = 0.023). This implies that a 300mg tafenoquine dose in a 60kg adult would result in an additional mean haemoglobin reduction at day 2 or 3 of around 0.1g/dL. Increasing the dose to 450mg would result in a 0.15g/dL reduction. In a separate model, day 7 MetHb concentration was correlated with haemoglobin reductions on day 2 or 3 (as postulated in the causal diagram in Figure 1). A one percent increase in MetHb was associated with a change in haemoglobin concentration of -0.08g/dL (95% CI: -0.12 to -0.03). In contrast, every one day increase in the tafenoquine terminal elimination half-life was associated with a haemoglobin increase of 0.02g/dL (95% CI: 0.01 to 0.04).

## Discussion

Our analysis provides strong evidence that the currently recommended adult dose of tafenoquine is insufficient for radical cure in all adults. In endemic areas, relapse of vivax malaria is a major contributor to morbidity and, in areas of high transmission, to mortality - particularly in young children [30, 35, 36]. Tafenoquine prevents vivax malaria relapses in a single dose treatment. It is potentially, therefore, a major advance in antimalarial therapeutics [3]. Suboptimal dosing severely reduces its therapeutic utility. Our pharmacometric assessment shows that the antimalarial dose-response relationship is steep around the currently recommended dose. Heavier patients are particularly disadvantaged. Substantial improvements in radical curative efficacy are possible with small dose increases. The tafenoquine dose-response relationship for haemolysis in G6PD deficiency is much less well characterised [10], although the currently recommended dose is considered potentially dangerous in all G6PD deficient individuals, including female heterozygotes who may still have a large proportion of G6PD deficient red cells. For these reasons substantial efforts have been made to ensure accurate quantitative testing so that tafenoquine is not given to G6PD deficient individuals. The efficacy, tolerability and safety of increased doses should be evaluated now in prospective studies. Given the substantial investment to date in developing and promoting tafenoquine, and the associated investments in developing field applicable methodologies for quantitative G6PD estimation, a relatively small further investment to find the correct dose would seem thoroughly worthwhile. Assuming a distribution of body weights similar to that in the tafenoquine pre-registration trials, we estimate that increasing the current dosing recommendation from 300mg to 450mg will more than halve recurrence at 4 months. This would be operationally feasible given the current 150mg tablet size. A 50% increase in the adult dose, resulting in at most 12.9mg/kg in a 35kg adult, is predicted to avert one relapse of malaria for every 11 patients treated.

The mechanism of the relapse preventing (radical curative) action of tafenoquine is still uncertain. This large individual patient data meta-analysis, which comprises data from the majority of all patients enrolled in tafenoquine clinical trials, shows clearly that the radical curative activity of tafenoquine in vivax malaria results from the activity of its oxidative metabolites. Dose, rather than exposure to the parent compound, was the main determinant of radical curative efficacy. Despite reducing total exposure, more rapid tafenoquine elimination, and therefore greater biotransformation, was associated with increased efficacy. This suggests, unsurprisingly, that tafenoquine behaves pharmacologically like a slowly eliminated form of primaquine [37]. The central importance of the oxidative metabolites in antimalarial activity was confirmed in these large and detailed studies by the relationship of radical curative efficacy with methaemoglobinaemia. A similar relationship has been shown for other 8-aminoquinolines [6, 15]. The reversible oxidation of intraerythrocytic haemoglobin reflects oxidative activity, and in this large series was correlated positively with relapse prevention and negatively with the tafenoquine half-life. Tafenoquine causes very minor oxidative haemolysis in G6PD normal individuals, and this too correlated with methaemoglobinaemia. These findings are all internally consistent with the hypothesis that oxidative activity is necessary for radical curative activity and also mediates haemolysis. Although tafenoquine has been engineered to be stable, it therefore does require metabolism for its pharmacological activity. For primaquine the unstable hydroxylated metabolites have been implicated as the active moieties, with CYP2D6 playing an important role in their formation [38]. The identity of the tafenoquine active oxidative metabolites, and the metabolic pathway have not been characterised yet. These data confirm the value of methaemoglobinaemia as a readily quantifiable non-invasive pharmacodynamic correlate of the antimalarial activity of the 8-aminoquinoline antimalarials [6, 15].

The inferred steep antimalarial dose-response relationship provides a satisfactory explanation for the poor recent results with tafenoquine reported in the INSPECTOR trial [12] from Indonesia. In this trial the currently recommended fixed dose of 300mg was given to a group of Indonesian soldiers who had been exposed to high malaria transmission. The soldiers were heavier than the Southeast Asian patients studied in the registration trials, with a mean body weight of 70kg. As Southeast Asian *P. vivax* infections are thought to require larger doses of primaquine than elsewhere, it is therefore likely that the high preceding transmission in Papua (heavier hypnozoite burdens) [14] and lower mg/kg dosing combined to result in very poor radical curative efficacy.

Our analysis was confined to assessment of the determinants of radical curative efficacy. The main limitations of the study are the absence of direct measurements of the bioactive metabolites of tafenoquine, and imprecision in the measurement of relapse. As relapses can be with parasites which are genetically unrelated to the primary infection, distinguishing recrudescence, relapse and reinfection is difficult. Combining genotyping with time to event analysis improves discrimination [39]. In these clinical trials parasite genotyping was only done at three microsatellite loci and raw genotyping data could not be obtained to infer probabilistic assessments of relapse versus reinfection [39]. However, these studies were conducted in generally low transmission settings, and nearly all recurrences occurred within four months of treatment which makes a significant contribution from reinfection unlikely (Figure 2). As tafenoquine metabolites could not be measured their production was inferred from the parent drug elimination rate. The pharmacodynamic properties of the oxidative metabolites were inferred from measurements of methaemoglobin blood concentrations. Methaemoglobin production does not lie on the causal pathway to the killing of hypnozoites, and measurements show substantial variation between patients, but it proved to be a useful, non-invasive, easily measured, and therefore readily deployable, surrogate for radical curative activity. The observed correlation of recurrence with terminal elimination half-life and MetHb production strongly supports the proposed causal model (Figure 1). Although the importance of drug metabolism was shown, our large study was insufficiently powered to evaluate the role in tafenoquine bioactivation of *CYP2D6* genetic polymorphisms (which have been shown to be important in bioactivating primaquine). Further work to characterise tafenoquine metabolism is needed.

Finally, only DETECTIVE part 1 randomised patients to different tafenoquine doses; in the other two studies, variation in mg/kg dose results from variation in patient weight. Therefore, some confounding because of population differences in recurrence risk for patients of different body weight (e.g. different socioeconomic status) cannot be excluded. Patient weight was not a predictor of recurrence per se. Over half the variation in body weight is polygenic, which is highly unlikely to be unrelated to malaria risk [40]. Importantly the relationship between dose and recurrence was consistent across all 3 geographic regions. The results are nearly identical when only analysing data from DETECTIVE part 1 (OR: 0.62 per mg/kg increase [95% CI 0.52 to 0.72]).

These well conducted, detailed pre-registration clinical studies provide a clear description of the clinical pharmacology of this new slowly eliminated 8-aminoquinoline in the radical cure of vivax malaria. The results show the central importance of tafenoquine metabolism to oxidative intermediates in determining therapeutic efficacy. They allow accurate characterisation of the tafenoquine antimalarial dose-response relationship, and thereby provide compelling evidence that the currently recommended adult dose is insufficient. In order to optimise the radical cure of vivax malaria with tafenoquine, prospective studies should now be instituted to assess the tolerability, safety and efficacy of higher doses. Increasing the adult dose to 450mg is predicted to substantially reduce the risk of relapse, particularly in heavier patients.

## Data Availability

https://github.com/jwatowatson/Tafenoquine-efficacy

## Acknowledgements

We thank GlaxoSmithKline Research & Development Ltd for providing access to the clinical trial data via the www.clinicalstudydatarequest.com website.

This work was supported by the Wellcome Trust. A CC BY or equivalent licence is applied to the author accepted manuscript arising from this submission, in accordance with the grant’s open access conditions. NJW is a principal research fellow funded by the Wellcome Trust (093956/Z/10/C). JAW is a Sir Henry Dale Fellow funded by the Wellcome Trust (223253/Z/21/Z). RJC is funded by an Australian National Health and Medical Research (NHMRC) Emerging Leader Investigator Grant (1194702). RNP is a Wellcome Trust Senior Fellow in Clinical Science (200909). JAS is funded by an Australian NHMRC Leadership Investigator Grant (1196068).

## Supplementary Figures and Tables

### Tafenoquine drug levels: data pre-processing

#### Phase 1 data

In the phase 1 study there were 860 drug level measurements in 72 patients. A total of 14 datapoints were below the lower limit of quantification (BLQ). The lower limit of quantification was 2 ng/ml. We removed one drug level which was a clear outlier from visual inspection, leading to a total of 859 levels in 72 patients in the analysis dataset.

#### Efficacy trials data

A total of 651 patients were randomised to tafenoquine in the three efficacy trials. One patient was excluded as they vomited but were not re-dosed. Out of the remaining 650 patient, drug measurement data were available from 649 patients. Three patients only had BLQ values and were removed from the analysis. 6 measurements were done in duplicate and the mean observed value was used (if one was BLQ then we imputed it as 1 ng/ml, i.e. half the lower limit of quantification). A total of 12 drug levels were removed after visual inspection of the individual profiles, leading to a total of 3640 measurements in 646 patients (101 BLQ).

### Comparison of major predictors of tafenoquine efficacy

We compared the predictive power of the tafenoquine mg/kg dose, the tafenoquine terminal elimination half-life, and the day 7 methaemoglobin concentration in a multivariable penalised logistic regression model (*n*=566 with complete data for all three predictors). Under this model, the mg/kg dose, the day 7 methaemoglobin and the terminal elimination half-life were all significantly associated with recurrence (Figure S16).

**Figure S1:**
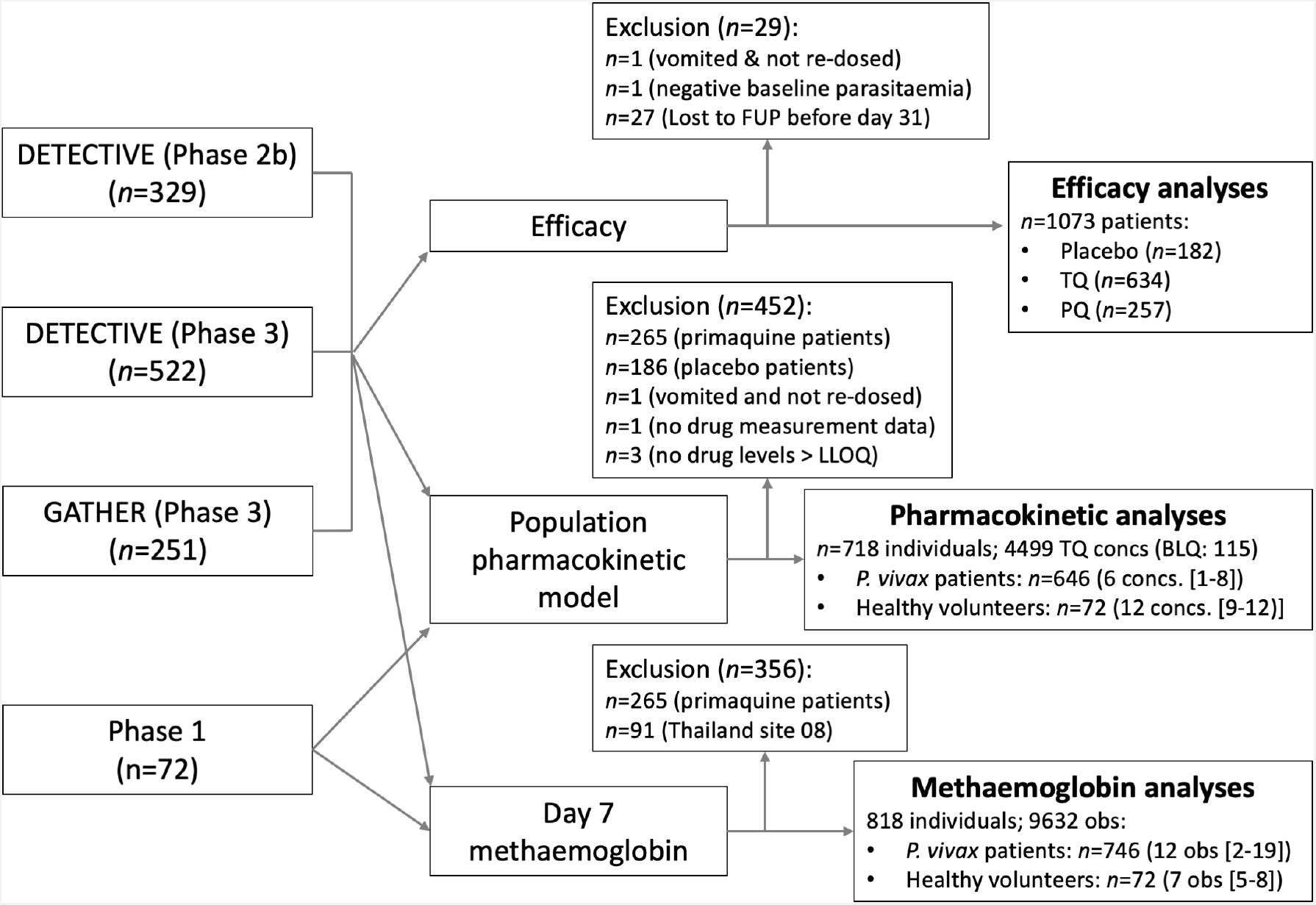
Flow diagram showing the construction of the analysis datasets for the binary end-point efficacy analyses (note that the patients who dropped out before day 31 are included in the time to event analyses), the pharmacometric analyses (AUC_[0,*∞*)_, C_max_, and *t*_1*/*2_) and the day 7 methaemoglobin analyses.

**Figure S2:**
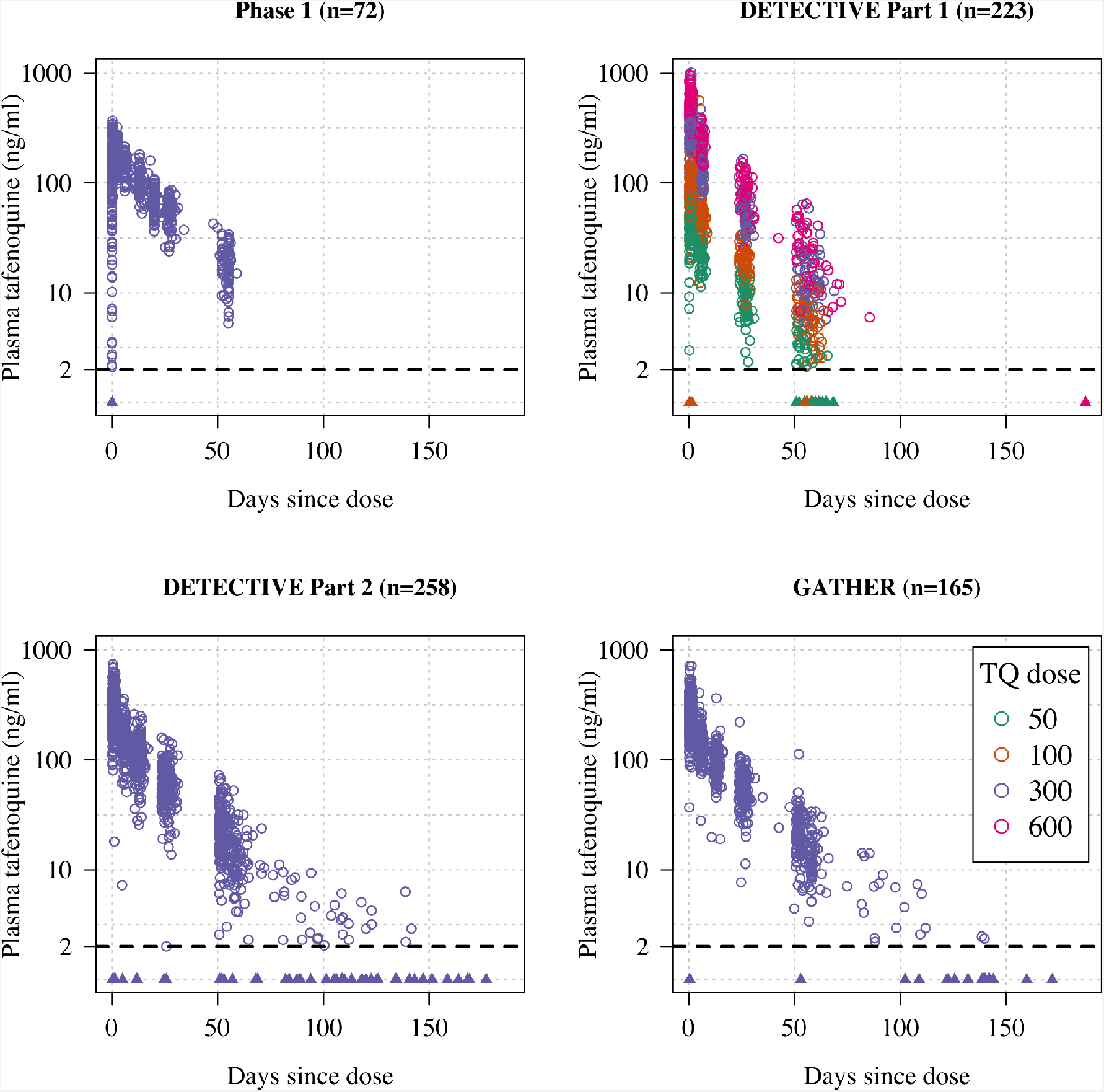
Plasma tafenoquine concentrations over time across the four studies (4499 concentrations in *n*=718 individuals). Concentrations below the lower level of quantification (BLQ) are shown by the triangles with a value of 1ng/ml (lower limit of quantification is shown by the dashed line at 2ng/ml). Circles are coloured by dose.

**Figure S3:**
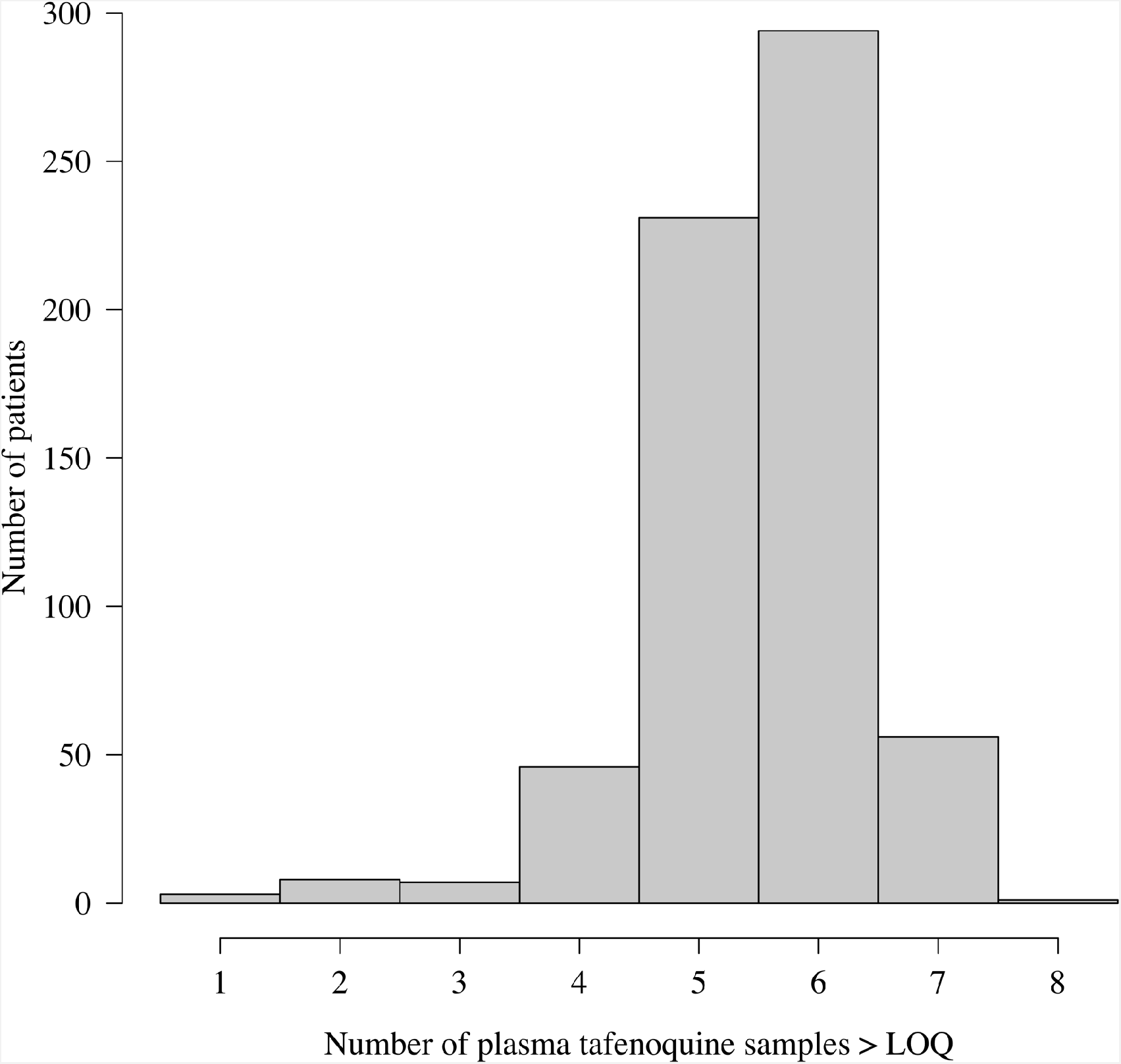
Distribution of the number of plasma tafenoquine concentrations per patient in the three efficacy trials. The very large majority of patients had 5 or 6 observed concentrations each. For those with few samples (e.g. 1-2), the estimates of the PK summary statistics (AUC_0,*∞*_, *C*_max_ and *t*_1*/*2_) will be shrunk towards the population mean estimates.

**Figure S4:**
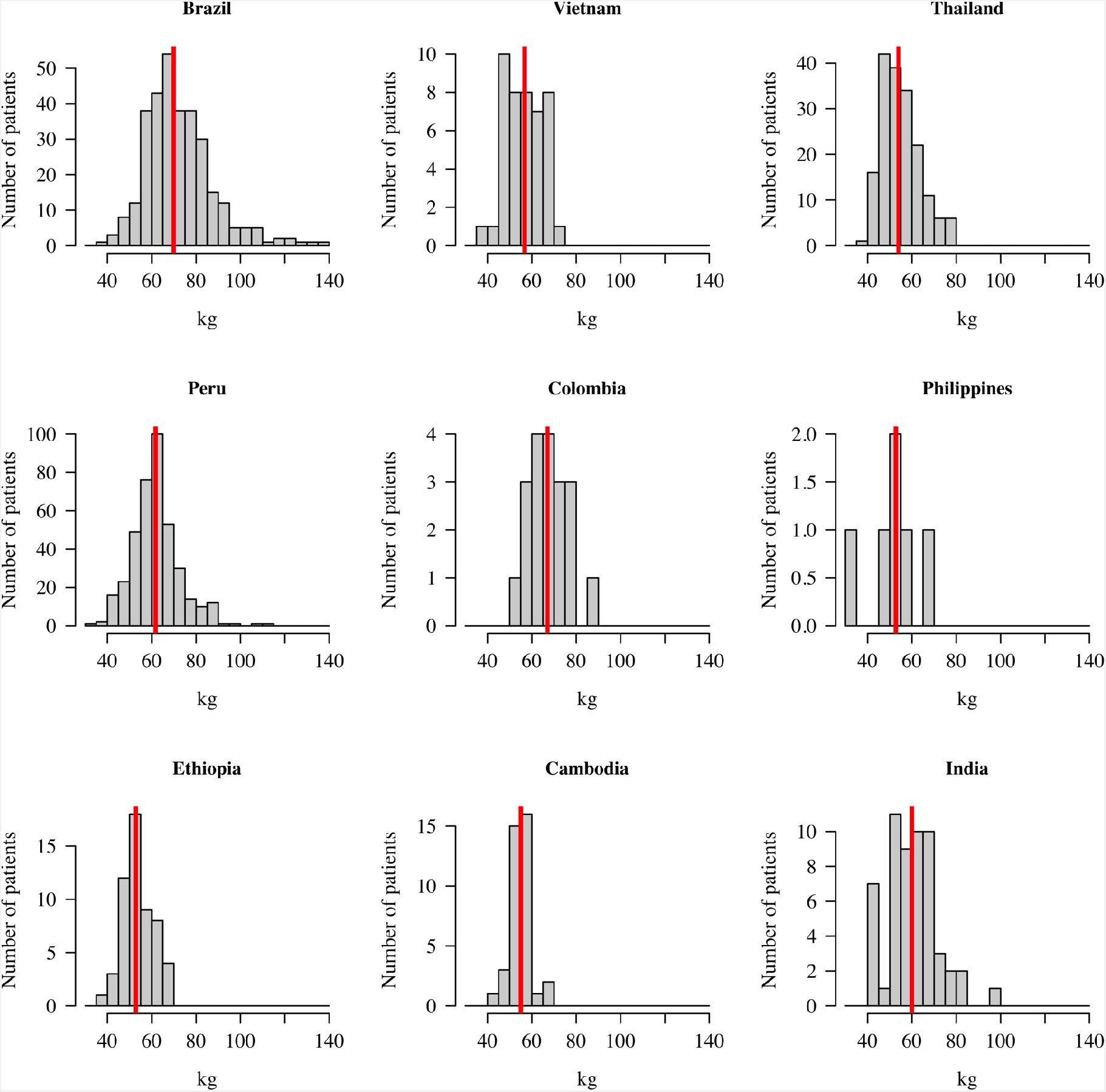
Distribution of patient weights across the nine countries. The vertical red lines show the median weights.

**Figure S5:**
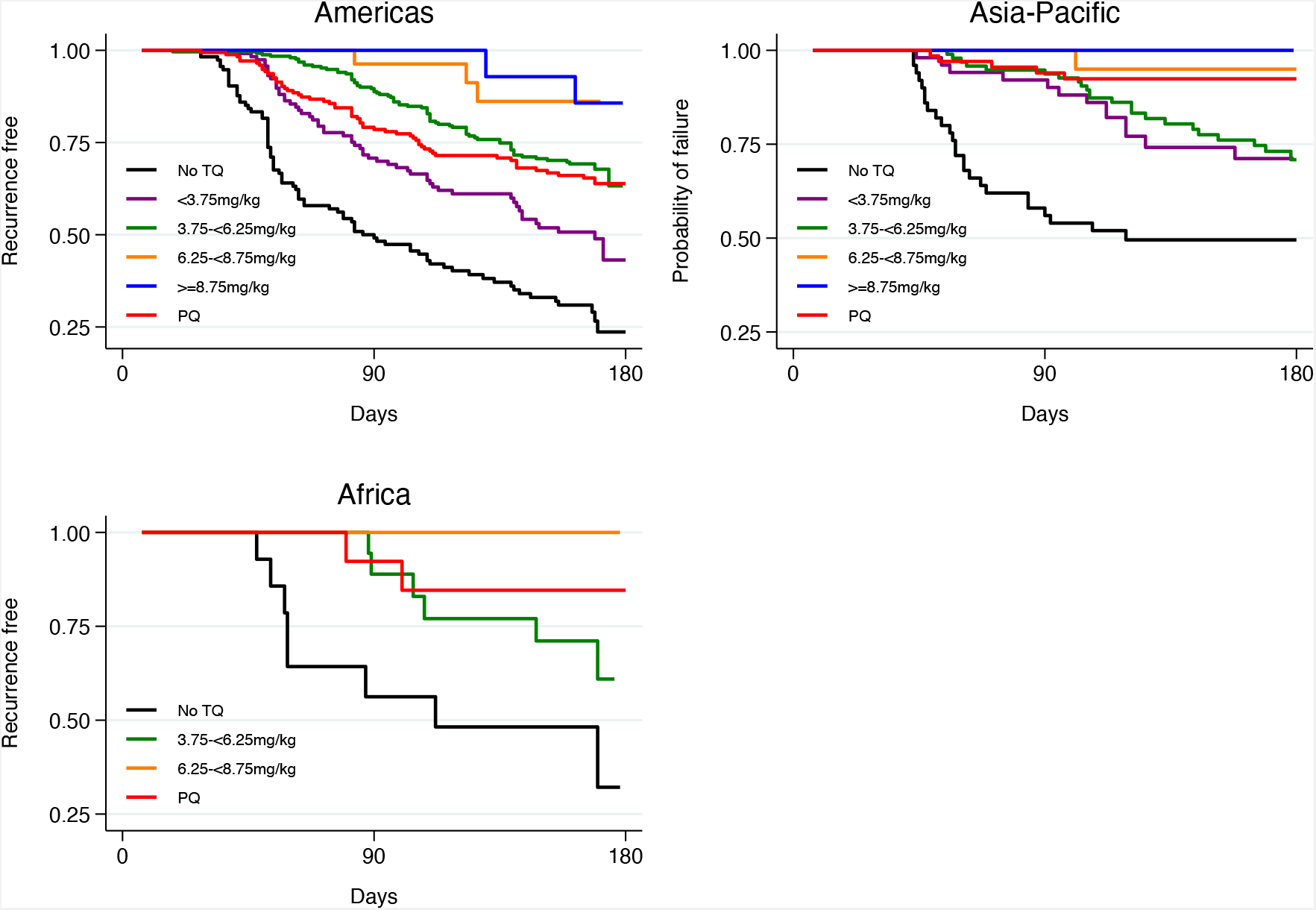
Kaplan-Meier survival curves for the time to first recurrence by region after administration of chloroquine without tafenoquine or with tafenoquine 300mg in *P. vivax* patients. Results include 455 patients from Americas, 171 patients from Asia-Pacific and 42 patients from Africa. TQ: tafenoquine.

**Figure S6:**
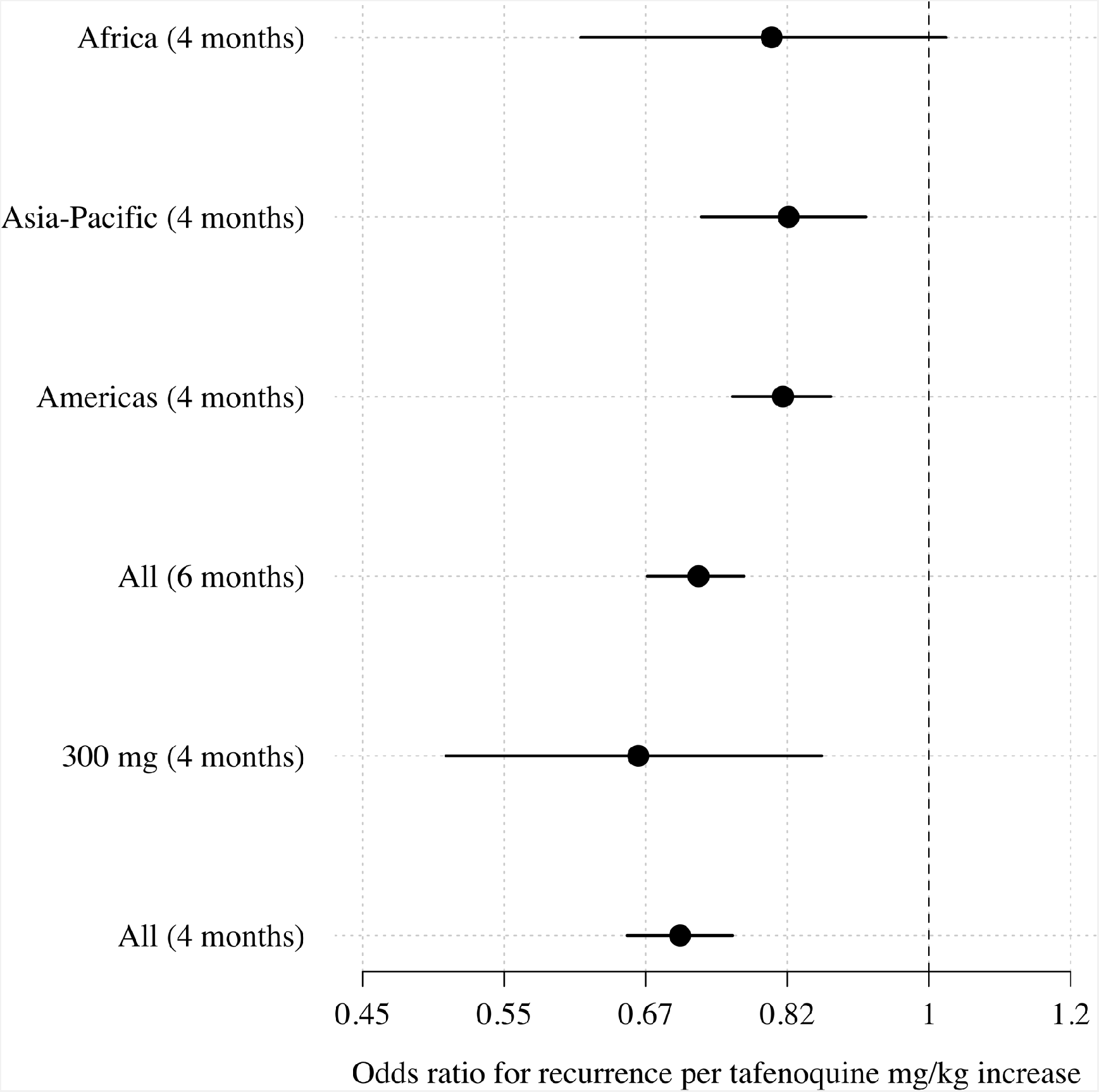
Comparison of estimated odds-ratios (95% CI) for the two endpoints (any recurrence by 4 months and any recurrence by 6 months); the sensitivity analysis in the 300mg group only; subgroups by geographic region.

**Figure S7:**
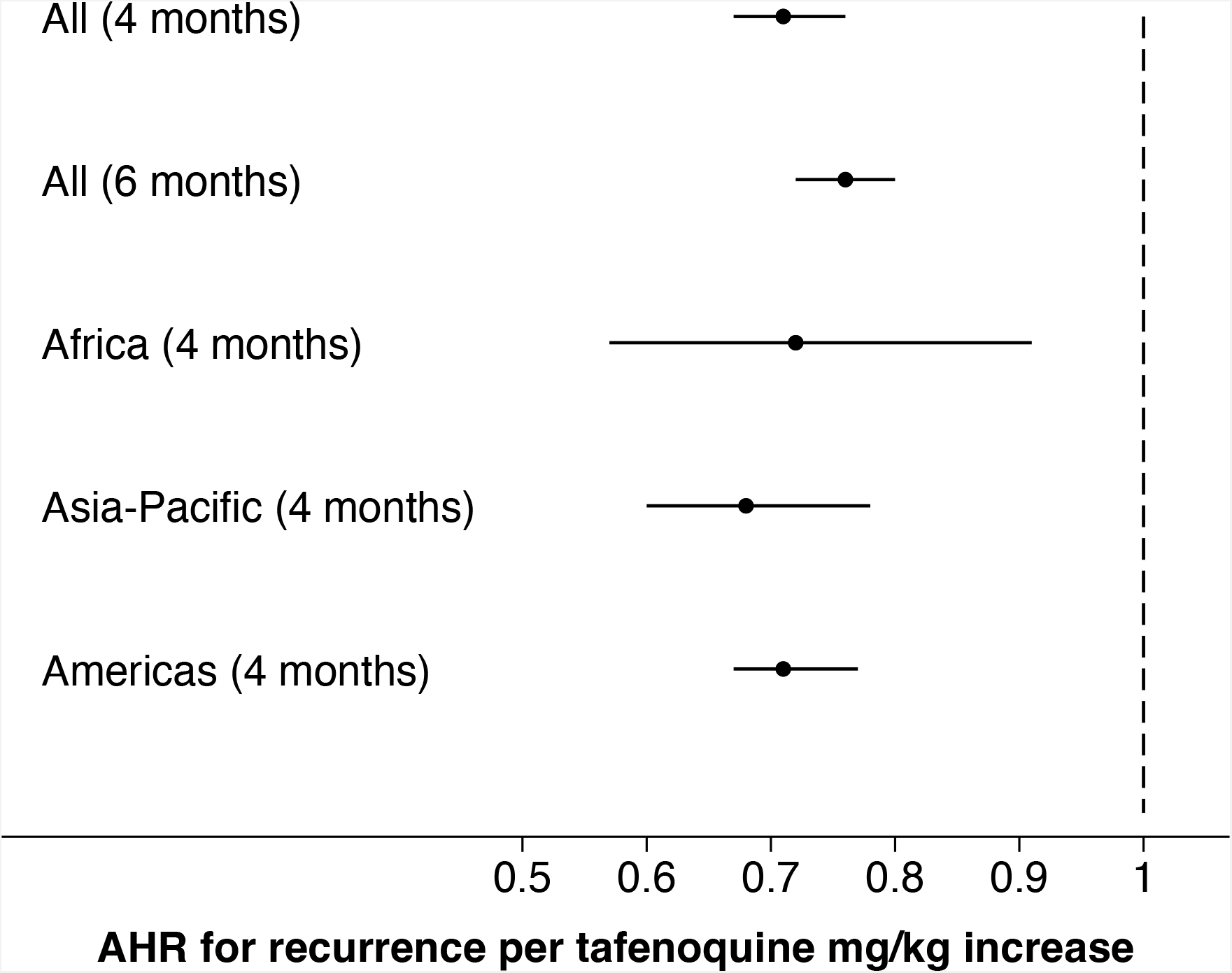
Comparison of estimated adjusted hazards ratio (95% CI) using time to event analysis for the two endpoints (any recurrence by 4 months and any recurrence by 6 months); subgroups by geographic region.

**Figure S8:**
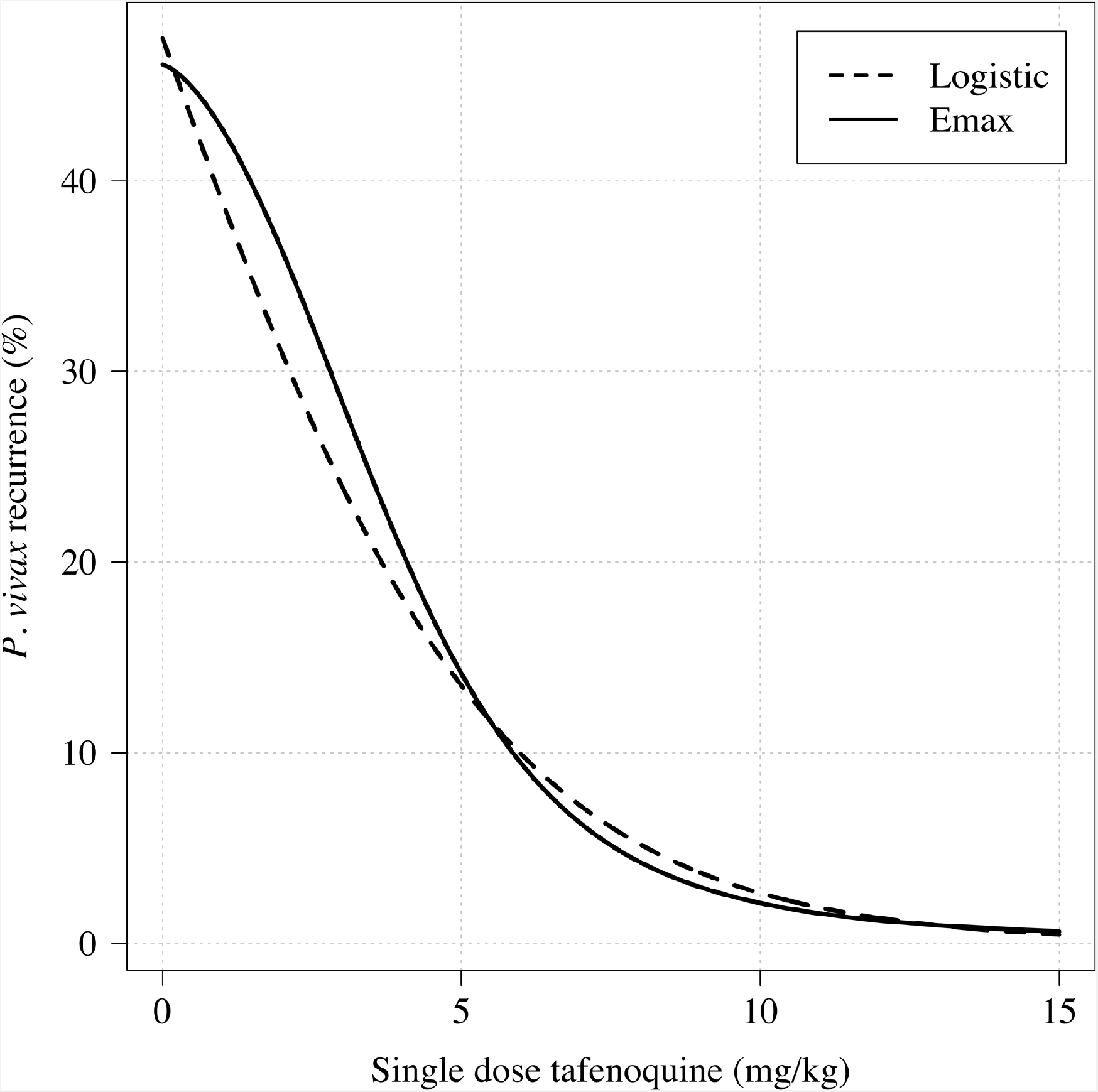
Comparison of logistic regression and E_max_ model fits for any recurrence within 4 months.

**Figure S9:**
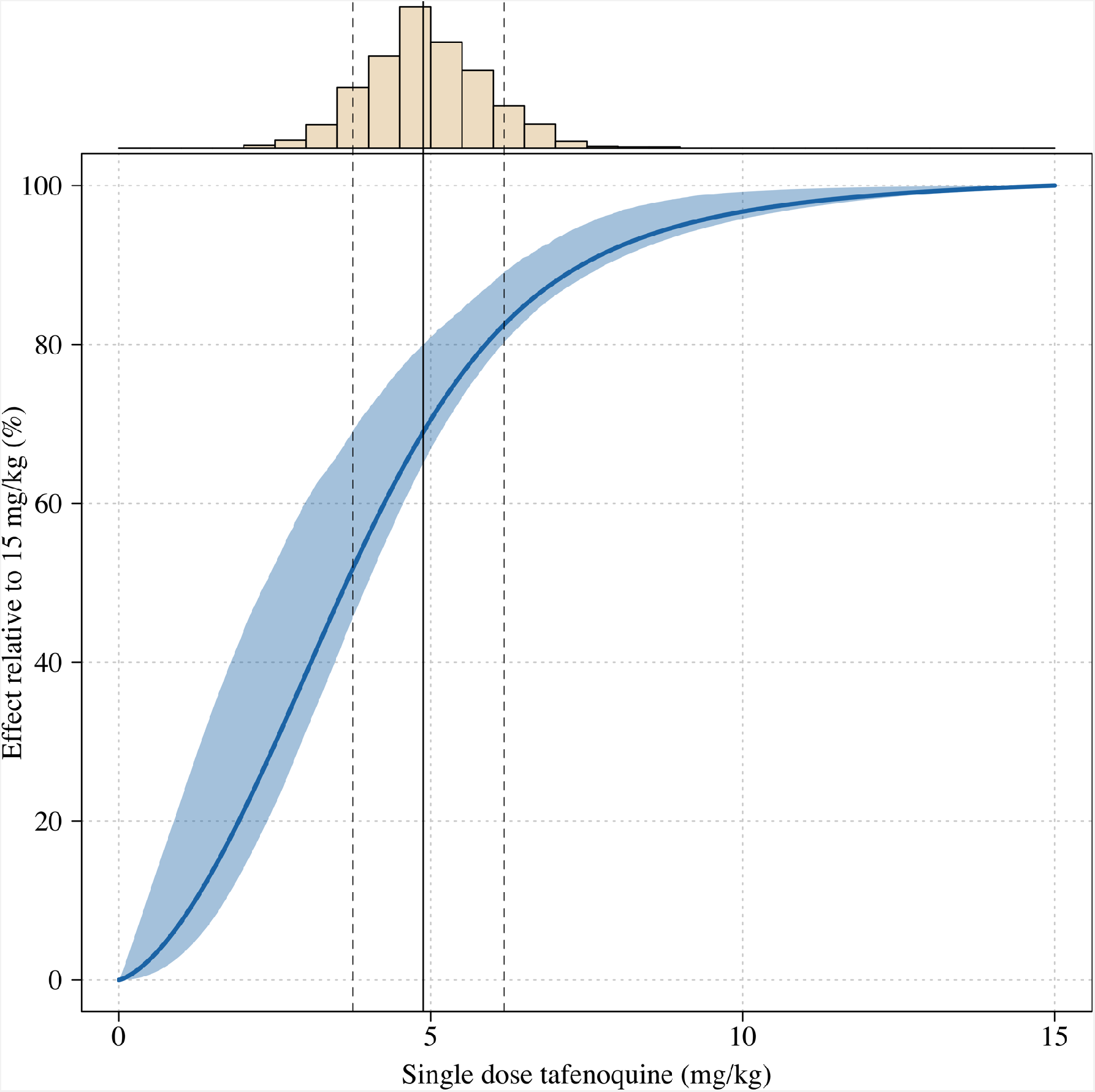
Estimated dose-response curve for tafenoquine mg/kg dose, whereby the maximal effect is defined as the effect for a 15mg/kg dose (the highest tolerated dose). The histogram above shows the distribution of mg/kg doses expected in patients using the empirical weight distribution from the three efficacy studies. The vertical lines show the 10, 50 and 90th percentiles of the expected mg/kg distribution.

**Figure S10:**
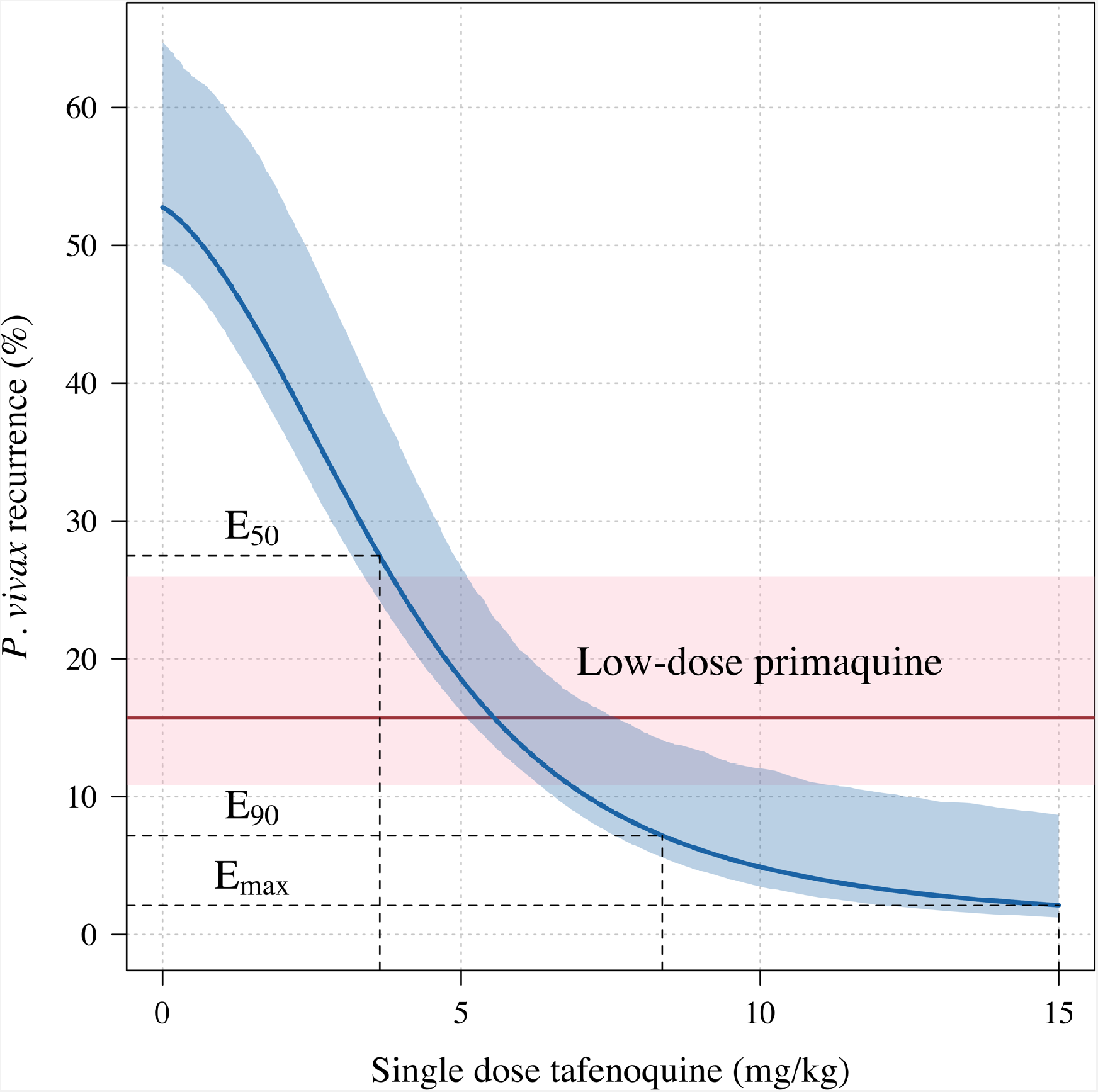
Tafenoquine mg/kg dose and the probability of any recurrence of *P. vivax* malaria at 6 months under the E_max_ model. Thick blue (shaded blue): E_max_ fit (95% CI) for any recurrence by 6 months; dashed red (shaded pink): estimated probability of any recurrence by 6 months after 3.5mg/kg primaquine (95% CI).

**Figure S11:**
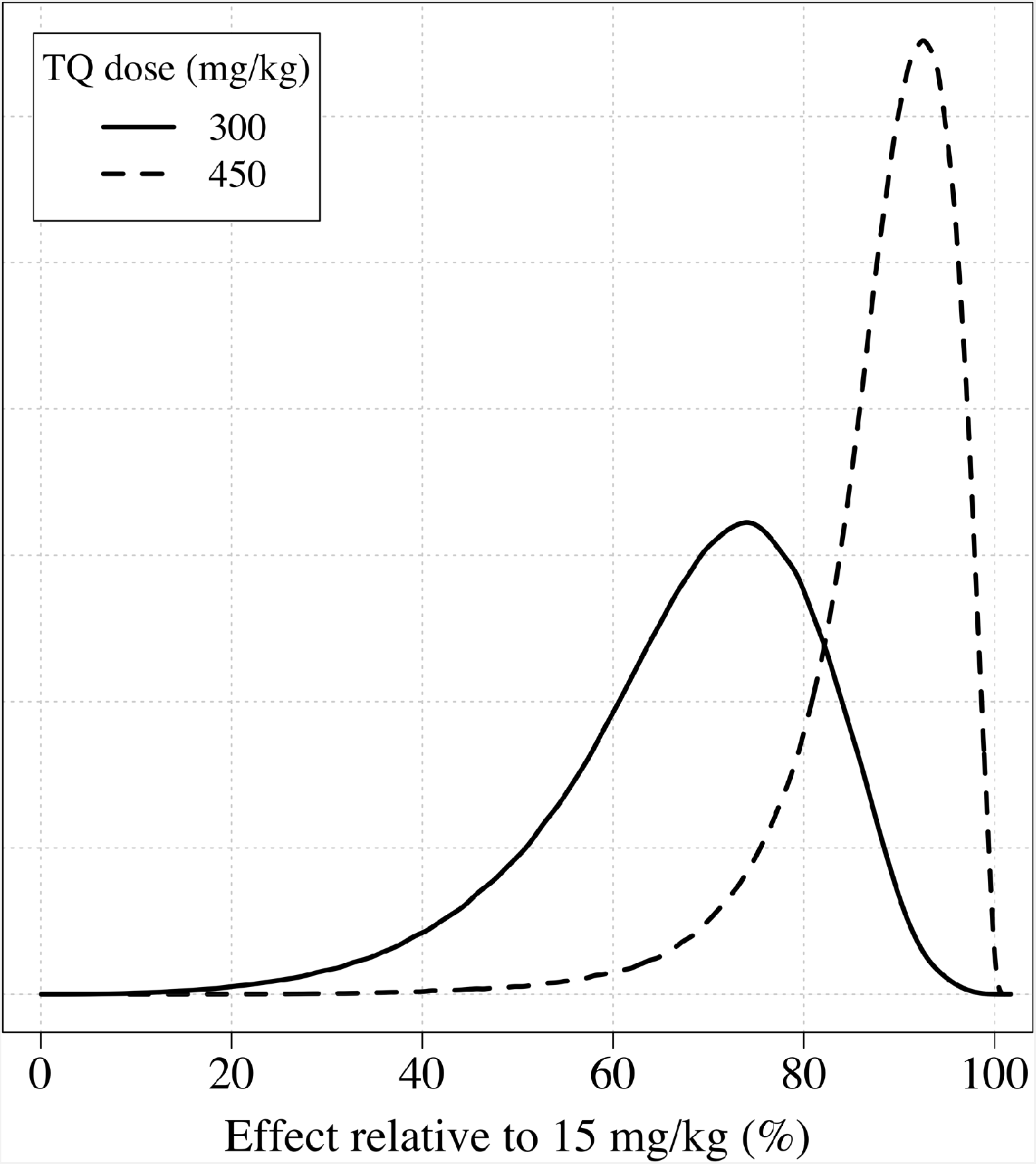
The distributions of the expected weight based efficacy (defined as the % reduction in recurrence at 4 months relative to the maximum dose of 15mg/kg) for a 300 mg single dose (thick line) and a 450mg fixed dose (dashed line). This is based on the empirical weight distribution from the three efficacy studies.

**Figure S12:**
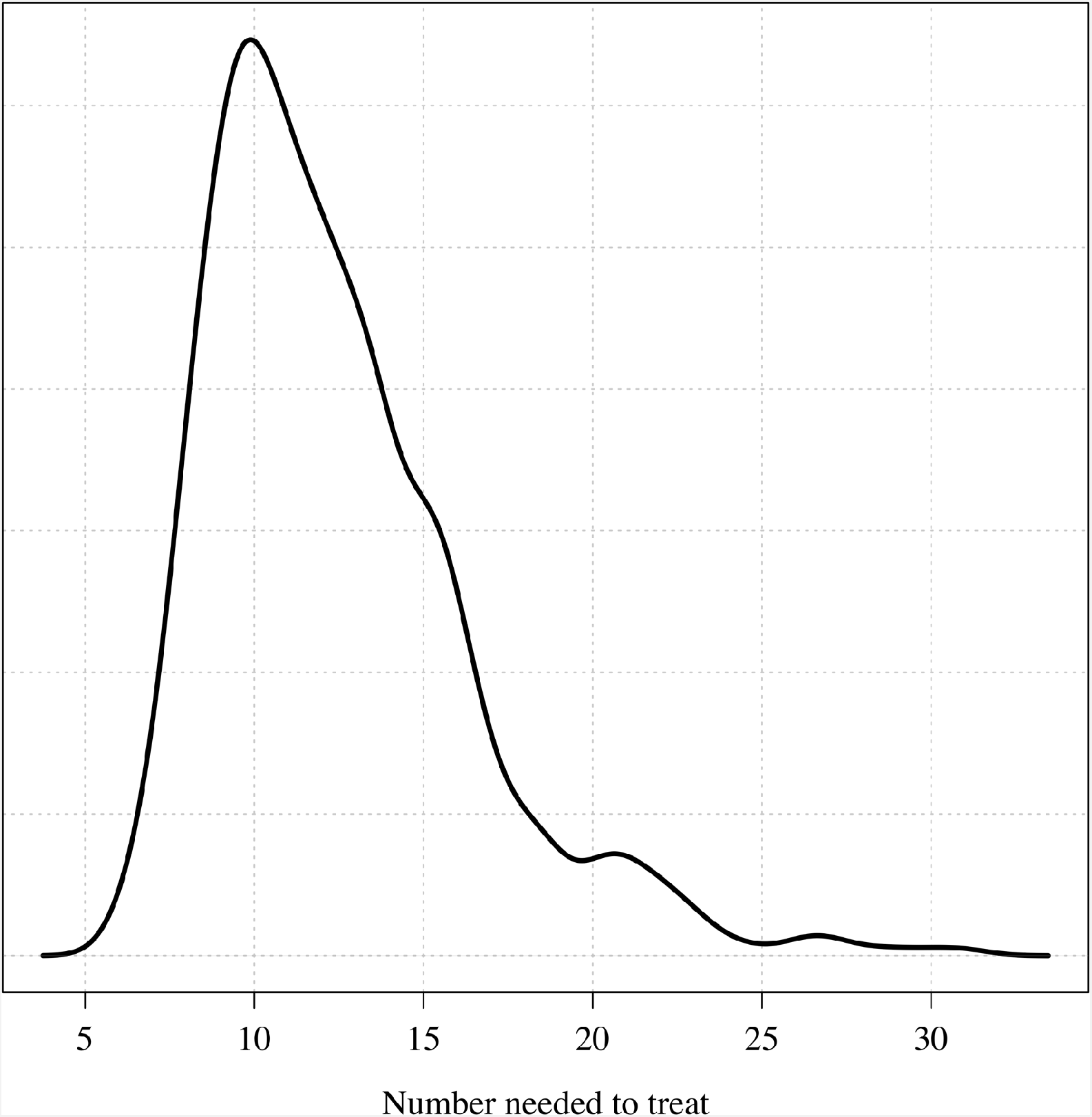
The posterior distribution over the number needed to treat to prevent one recurrence at 4 months under the Bayesian Emax model.

**Figure S13:**
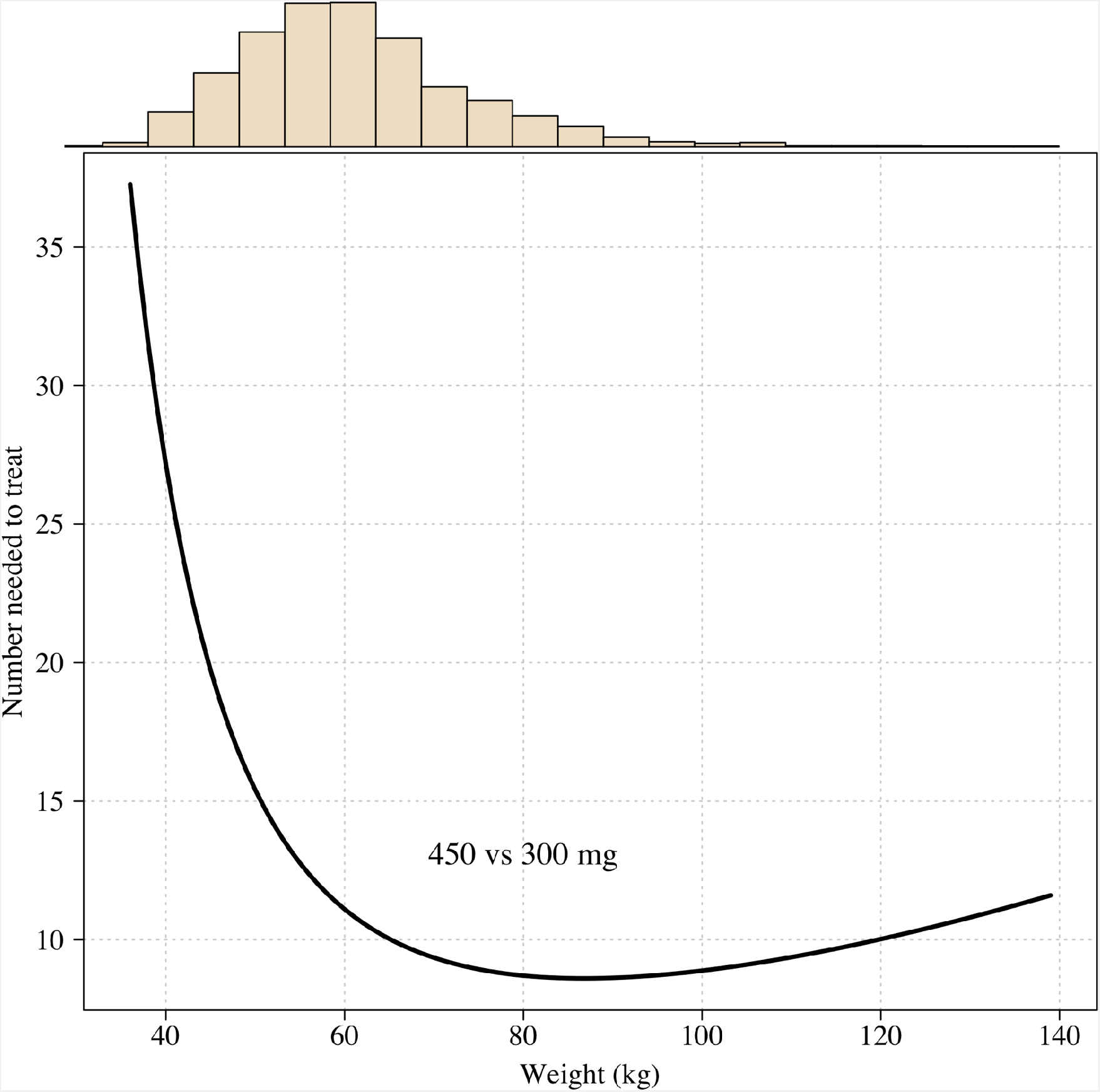
The number needed to treat with 450mg instead of 300mg to prevent one recurrence at 4 months as a function of patient weight under the Emax model.

**Figure S14:**
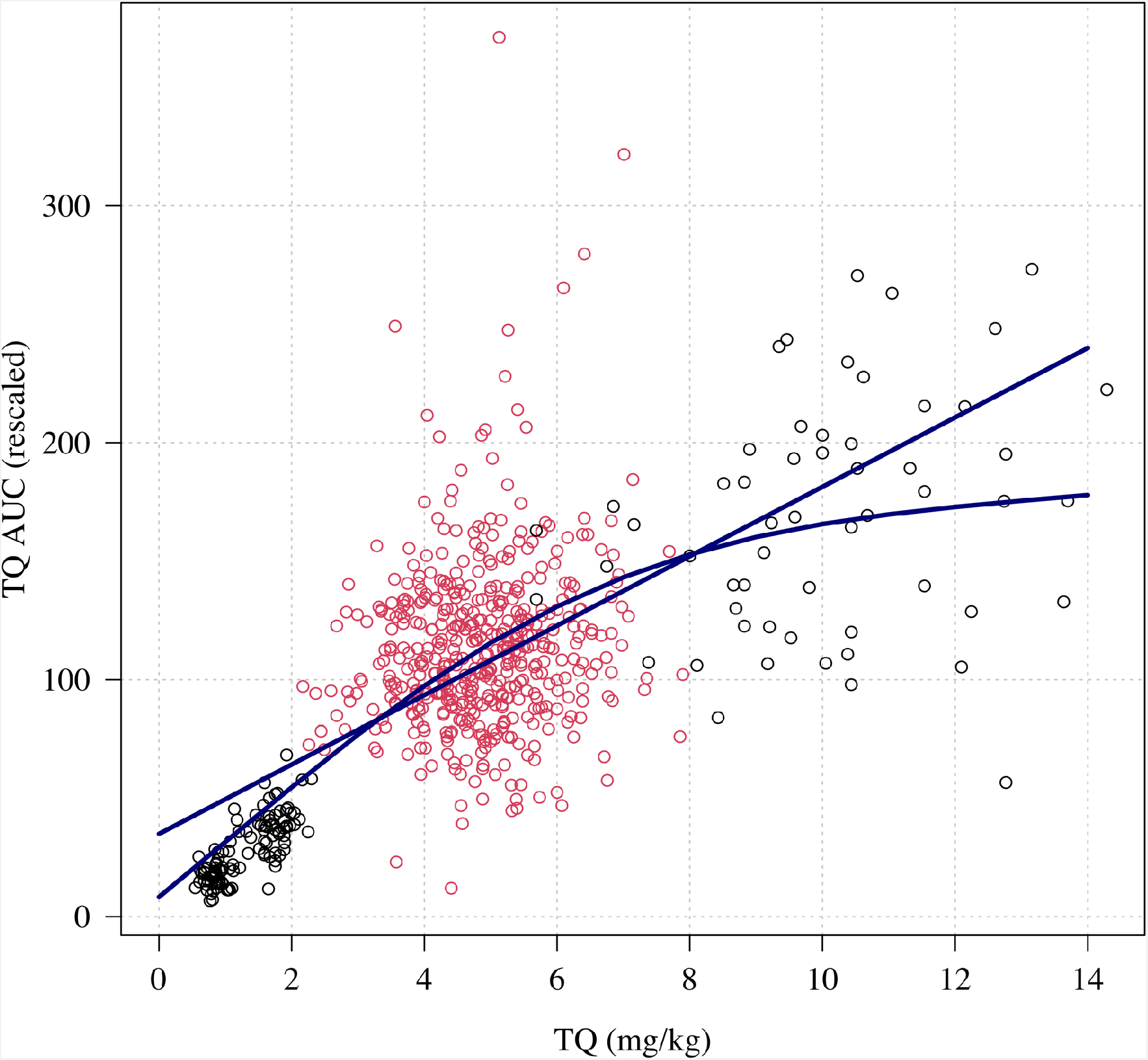
Relationship between mg/kg dose of tafenoquine and the AUC_[0,*∞*)_. Linear and spline fits are overlaid to show trend. The red dots show the patients who received a 300 mg dose (current recommended treatment), the black circles show the other doses (50, 100 and 600mg).

**Figure S15:**
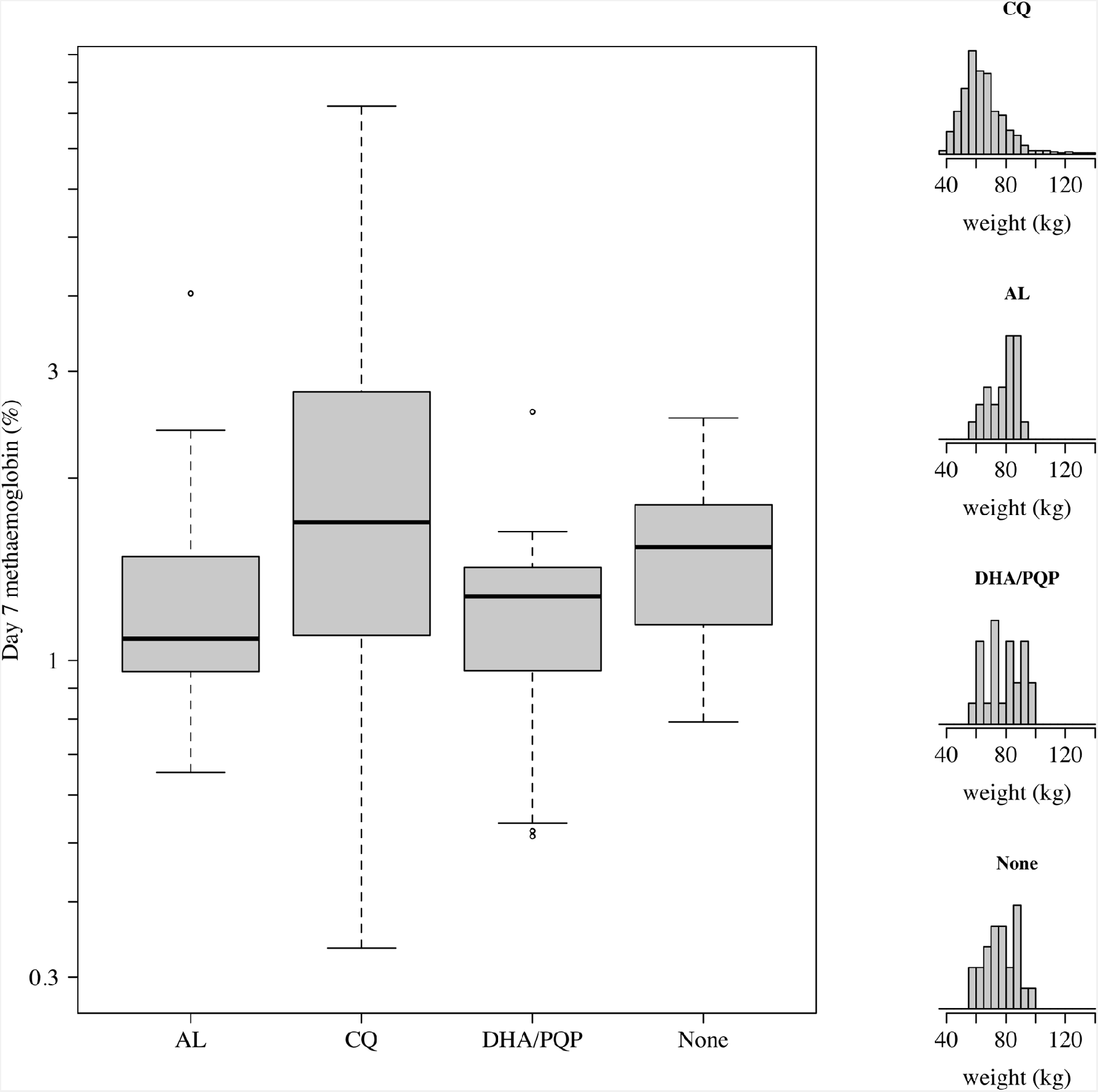
Comparison of day 7 methaemoglobin values after administration of tafenoquine 300 mg in *P. vivax* patients (all received chloroquine [CQ]) and healthy volunteers who were randomised to either no partner drug, AL, or DHA-PQP. The distributions of weights are shown in the right panels as the mg/kg dose is the primary driver of day 7 methaemoglobin. The mean weight in patients was slightly lower than in the healthy volunteers (although with a much larger variation).

**Figure S16:**
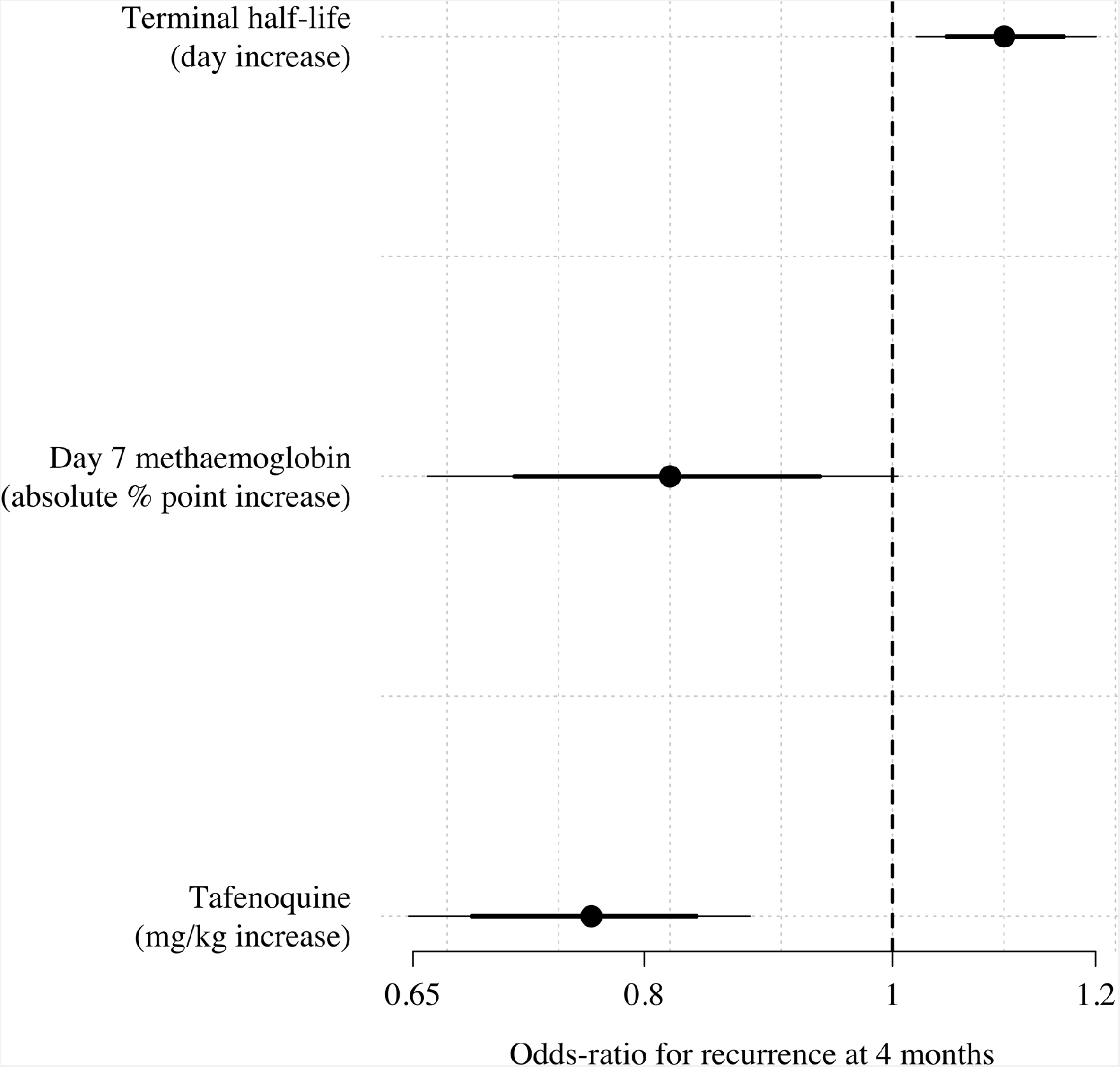
Multivariable model comparison of predictors of recurrence at 4 months in patients who received tafenoquine and who had recorded values (*n*=566). The circles (thick and thin lines) show the odds-ratios (80% and 95% credible intervals) for recurrence under a Bayesian penalised logistic regression model.

**Table S1:** Haematology safety outcomes by treatment arm. n (%); mean (sd)

| Outcome | No TQ | <3.75<br>mg/kg | 3.75-<br><6.25<br>mg/kg | 6.25-<br><8.75<br>mg/kg | >=8.75<br>mg/kg |
| --- | --- | --- | --- | --- | --- |
|  | n=186 | n=171 | n=378 | n=56 | n=44 |
| Hb fall >25% to <7<br>g/dL | 0 (0%) | 0 (0%) | 0 (0%) | 0 (0%) | 0 (0%) |
| Absolute Hb fall >5<br>g/dL | 0 (0%) | 0 (0%) | 1 (0.3%) | 0 (0%) | 0 (0%) |
| Hb fall to <5 g/dL | 0 (0%) | 0 (0%) | 0 (0%) | 0 (0%) | 0 (0%) |
| Hb change to day<br>2/3 | -0.4 (0.9) | -0.6 (0.9) | -0.7 (0.9) | -0.8 (0.8) | -0.6 (1.1) |
| Hb change to day<br>7±1 | -0.3 (0.9) | -0.3 (0.9) | -0.4 (1.1) | -0.4 (1.0) | -0.3 (1.3) |

